# Longitudinal microneedle sampling resolves tissue-restricted immune-microbiome dynamics in skin

**DOI:** 10.64898/2026.04.24.26351513

**Authors:** Ashok Kumar Dhinakaran, Anita Yvonne Voigt, Arthur Szacik, Soo Yeon Kang, Sebastian Giarratana, Julia Oh, Sasan Jalili

## Abstract

The skin microbiome shapes local immunity, but the mechanisms of microbiome-immune crosstalk remain poorly understood. A major barrier to discovery is the lack of approaches that enable simultaneous, longitudinal measurement of microbes and immune cells from the same tissue without disrupting barrier integrity. Here we present a hydrogel-coated microneedle (MN) patch that enables minimally invasive co-sampling of viable microbes, immune cells, and interstitial fluid from skin. In humans, the patches were well tolerated and preserved inter-individual microbial signatures. Murine models colonized with commensal *Staphylococcus epidermidis* and the opportunistic pathogen *Staphylococcus aureus*, revealed distinct immune trajectories during commensal colonization, pathogen challenge, and commensal-pathogen co-colonization. Pathogen colonization drives progressive inflammatory amplification, whereas commensal exposure induces controlled immune activation that stabilizes over time. Notably, *S. epidermidis* reshapes pathogen-induced responses, producing a transient immune activation followed by attenuation of inflammation. These results establish MN sampling as a strategy to resolve immune-microbiome dynamics in barrier tissues and provide a framework for mechanistic studies of host-microbe interactions in health and disease.

---

The human microbiome has emerged as a critical determinant of human health and disease, profoundly influencing immune function, barrier integrity, and resistance to pathogenic colonization^1,2^. As the largest organ and primary interface with the environment, skin is colonized by a complex microbial ecosystem exhibiting remarkable topographical diversity shaped by local physiological conditions^1,3,4^. The cutaneous microbiome is not merely a passive inhabitant; it actively maintains homeostasis through immune education and pathogen resistance via interactions with keratinocytes and immune cells^1,5–7^. However, systemic or local immune dysregulation and metabolic perturbations can disrupt this balance, triggering dysbiosis characterized by opportunistic pathogen expansion and decreased bacterial diversity^1,6^. This microbial imbalance plays a causal role in a range of cutaneous disorders such as atopic dermatitis and psoriasis, where specific dysbiotic signatures correlate with disease severity ^1,8,9^. The mechanistic links between systemic immune alterations, localized dysbiosis, and cutaneous inflammation remains incomplete, necessitating integrated approaches to examine host-microbiome interactions at multiple biological scales.

Beyond descriptive associations, a central unresolved question in cutaneous biology is how distinct microbial communities actively shape local immune trajectories in situ. Commensal organisms such as coagulasel⍰negative staphylococci can promote tissue-resident memory T cell formation, antimicrobial peptide production, and barrier reinforcement, thereby calibrating immune tone toward protective symbiosis^10,11^. In contrast, opportunistic pathogens such as *Staphylococcus aureus* trigger neutrophilic inflammation, Th17 polarization, and barrier disruption, often amplifying pre-existing immune dysregulation^12^. Critically, these divergent immune outcomes can arise within the same anatomical site and over short timescales, reflecting context-dependent interactions between microbial strain composition and local immune circuits. Dissecting these dynamics requires methods that capture microbial and immune states simultaneously and longitudinally from the same tissue microenvironment. Sampling microbiota in isolation cannot resolve whether observed dysbiosis is causal, consequential, or compensatory, and immune profiling alone cannot determine which microbial configurations drive protective versus pathogenic responses.

Microneedle (MN) patches are a minimally invasive platform capable of sampling both interstitial fluid (ISF) and immune cells without disrupting the local tissue microenvironment^13^. Unlike conventional surface sampling methods, MN technology penetrates the stratum corneum to reach the viable epidermis and upper dermis, enabling depth-controlled sampling at precise tissue layers where microbial communities and immune cells reside. By coating MN projections with cross-linked biocompatible polymers such as hydrogels that swell upon skin insertion, a porous matrix forms in situ to facilitate leukocyte infiltration and ISF extraction, allowing parallel monitoring of both cellular and soluble immune mediators^14,15^. Our group recently demonstrated the utility of hydrogel-coated MN patches for longitudinal immune monitoring, capturing tissue-resident memory T cells and cytokine profiles from skin with minimal tissue perturbation^16^4/22/2026 4:11:00 PM. While MN platforms have been explored for bacterial detection in food samples using bacteriophage loading^17^ or for pathogen sensing via silk fibroin arrays^13^, no study has demonstrated the use of MN patches to directly sample the native human skin microbiome in combination with immune sampling.

Here, we introduce a hydrogel-coated MN patch that enables simultaneous sampling of viable skin microbiota and immune cells from deep tissue layers in a single, minimally invasive intervention. *In vitro* studies confirmed efficient bacterial capture on the hydrogel surface, including SEM visualization of bacterial adhesion. We then demonstrate that this platform can reliably recover native skin microbiota from both mouse skin and healthy human subjects, with successful sampling achieved after short (20 min) and extended (24 h) application times. We use murine *S. aureus* skin colonization and infection models to longitudinally profile bacterial dynamics together with infiltrating immune cell populations at defined sites over time. MN patches applied to acute (1-day) and chronic (3-day) infection sites yielded superior bacterial recovery relative to conventional swabbing or skin-digest approaches, while simultaneously capturing immune cells recruited to infected tissue. These data establish our MN platform as a uniquely powerful tool for integrated, minimally invasive monitoring of microbial communities and immune responses in both health and disease, and provide a foundation for dissecting how commensals and pathogens dynamically shape cutaneous immunity *in vivo*.

## Results

### MN platform enables multimodal and unbiased microbial sampling of the cutaneous microenvironment

To enable longitudinal interrogation of immune-microbiome dynamics, we leveraged a minimally invasive MN patch platform capable of simultaneously sampling ISF, viable immune cells, soluble mediators, and microbial communities from human and murine skin. Unlike conventional surface swabs, which primarily access the stratum corneum, or biopsies, which are invasive and unsuitable for repeated sampling, our MN arrays penetrate the viable epidermis to capture a spatially integrated snapshot of the local cutaneous microenvironment (**Fig. 1a**). Each patch consists of 400 square-pyramidal poly(L-lactic acid) (PLLA) MN projections (600□μm height, 300□μm base width) fabricated by melt-molding, providing sufficient mechanical strength for reproducible epidermal insertion while remaining minimally invasive (**Fig. 1b**). The MN arrays are coated with a thin hydrogel sampling layer composed of ultrapure alginate solution and sucrose, which is ionically crosslinked with calcium chloride to form a conformal coating over each projection (**Fig. 1b**; **Supplementary Fig.1a**). Macroscopic and scanning electron microscopy imaging confirmed structural uniformity and preservation of sharp needle geometry following hydrogel coating (**Supplementary Fig. S1b**). Upon application with mild pressure, the alginate layer rapidly hydrates and swells, absorbing ISF and capturing components of the local skin microenvironment, including immune cells and microbial taxa present within and beneath the stratum corneum. All materials used in patch fabrication are Generally Recognized as Safe by the U.S. FDA, supporting their translational applicability.

**Figure 1.**
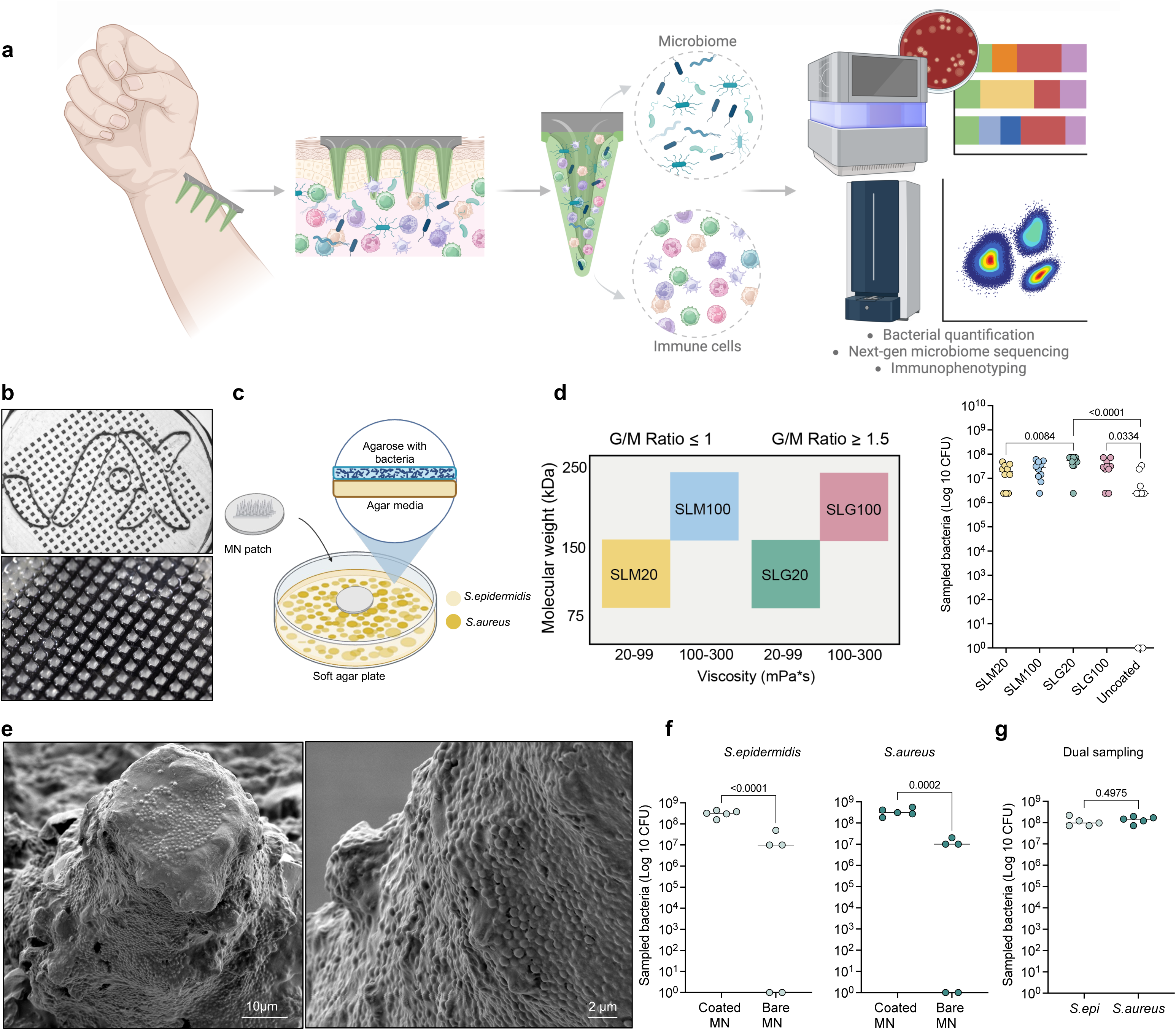
Hydrogel-coated microneedle patches enable multimodal and unbiased microbial sampling. **a**, Schematic view of the multimodal sampling approach of microbiome and immune cells using the microneedle patch. **b**, photographs of the hydrogel-coated MN patch. **c**, Schematic view showing the *in vitro* bacterial sampling from a soft agar plate using the MN patches. **d**, Molecular weight, G/M ratio and viscosity of different types of alginates tested as coatings for the MN patches and comparison of bacterial sampling capacity between patches coated with different alginates (n=10 patches per group). **e**, Scanning electron micrograph of the patches after 20 minutes *in vitro* sampling of the bacteria showing *S. aureus* sampled and retained on the hydrogel coating of the patch. **f**, Quantification of the *S. aureus* and *S. epidermidis* CFU counts for single strain sampling by the patches. **g**, Quantification of the *S. aureus* and *S. epidermidis* CFU counts for multi-strain sampling by the patches. Data shown are mean±□SEM. P values were determined using one-way ANOVA followed by Tukey’s multiple comparisons test (d,f,g). Panels created in BioRender: a,c.

To optimize microbial sampling, we evaluated whether alginate physicochemical properties influence bacterial capture efficiency. Using soft agar plates seeded with *Staphylococcus epidermidis* and/or *S. aureus*, two clinically relevant skin commensal and pathogenic species, MN patches were gently inserted into soft agar plates for 20 minutes, followed by enzymatic decrosslinking and colony-forming unit (CFU) quantification (**Fig. 1c**). We compared four alginate variants differing in molecular weight, viscosity, and guluronic-to-mannuronic acid (G/M) ratios (**Fig. 1d**). MN patches coated with SLG-20 alginate demonstrated significantly enhanced bacterial sampling compared to other alginate compositions, recovering higher *S. aureus* CFU numbers (**Fig. 1d**). We thus focused further studies on SLG20 as the sampling layer polymer. Scanning electron microscopy of individual SLG-20 MN projections confirmed dense bacterial adherence to the hydrogel-coated projection (**Fig. 1e**), validating effective microbial entrapment. Importantly, MN patches recovered comparable CFU levels when sampling mono-species cultures of *S. epidermidis* or *S. aureus* (**Fig. 1f**), and dual-species plates yielded balanced recovery of both taxa (**Fig. 1g**). These findings indicate that hydrogel composition critically influences sampling sensitivity while enabling simultaneous, unbiased capture of multiple bacterial species tested here, supporting its suitability for downstream microbiome profiling.

Because microbial viability is critical for accurate microbiome profiling, we evaluated whether alginate encapsulation and the enzymatic decrosslinking process affected bacterial survival. Fluorescence imaging demonstrated robust bacterial viability within the alginate layer before and after incubation (**Supplementary Fig. S1c,d**). Consistently, CFU quantification revealed no significant difference between untreated bacteria and bacteria recovered following alginate encapsulation and alginate lyase-mediated release (**Supplementary Fig. S1e,f**), indicating that the hydrogel coating and retrieval process are non-toxic and do not impair recovery of the Staphylococcus species examined. To determine whether microbial capture depends on penetration depth, we performed controlled sampling experiments using low-melting agarose plates in which *S. aureus* was embedded approximately 700 μm beneath the surface (**Supplementary Fig. S1g**). Longer microneedles (1000 μm) efficiently recovered bacteria from the embedded layer, whereas shorter microneedles (600 μm) failed to do so (**Supplementary Fig. S1h,i**), demonstrating statistically significant depth-dependent sampling. These results confirm that microbial recovery is governed by mechanical penetration depth rather than passive surface adsorption, supporting the capacity of MN arrays to access spatially distinct epidermal niches.

### MN sampling recapitulates murine skin microbial composition

MN patches were applied on the skin of C57BL/BJ mice for either 20 minutes or 24 hours, and microbiome profiles were compared with matched skin swabs and tissue biopsies using shotgun metagenomic sequencing (WGS) (**Fig. 2a**). At phylum level, Bacteriodota (formerly Bacteroidetes) and Firmicutes were the most dominant across sample types (**Fig. 2b**, **Supplementary Fig. S2a**), consistent with established murine skin microbiome datasets. 20-minute MN applications yielded variable profiles (**Supplementary Fig. S2a**), likely reflecting the relatively low microbial biomass and dense hair architecture, which may amplify stochastic effects during short-duration sampling. In contrast, 24-hour application reduced variability in mice; 24-hour MN sampling reproducibly recovered taxa comparable to swabs, with broader dispersion in alpha and beta diversity metrics (**Fig. 2c,d; Supplementary Fig. S2b**), suggesting that longer MN sampling captures a broader representation of the cutaneous microbial community beyond the stratum corneum. Swabs captured the largest median diversity, whereas biopsies showed enrichment for *Staphylococcus xylosus*, suggesting sampling biases associated with tissue excision and localized microbial distribution. Notably, 24-hour MN patches captured increased representation of Firmicutes and occasionally *Lactobacillus johnsonii*, a gut-associated microbe, suggesting access to microbial niches not readily captured by surface swabbing (**Supplementary Fig. S2c,d**). Across animals, inter-sample variation was comparable between 20-minute MN and swab modalities (**Fig. 2c,d; Supplementary Fig. S2**), indicating that biological variability exceeded modality-driven effects. *De novo* metagenomic assemblies largely recapitulated taxonomic assignments obtained via MetaPhlAn4 without substantially increasing resolution, supporting robustness of taxonomic classification. Reads corresponding to Flavobacterium were excluded from downstream analyses after identification as originating from the alginate lyase digestion reagent, consistent with known reagent-associated contaminants^18^. Together, these findings suggest that extended MN application may facilitate access to ecologically distinct compartments of the murine skin microbiome, given their capture of species represented both by surface skin swabs as well as gold standard biopsies.

**Figure 2.**
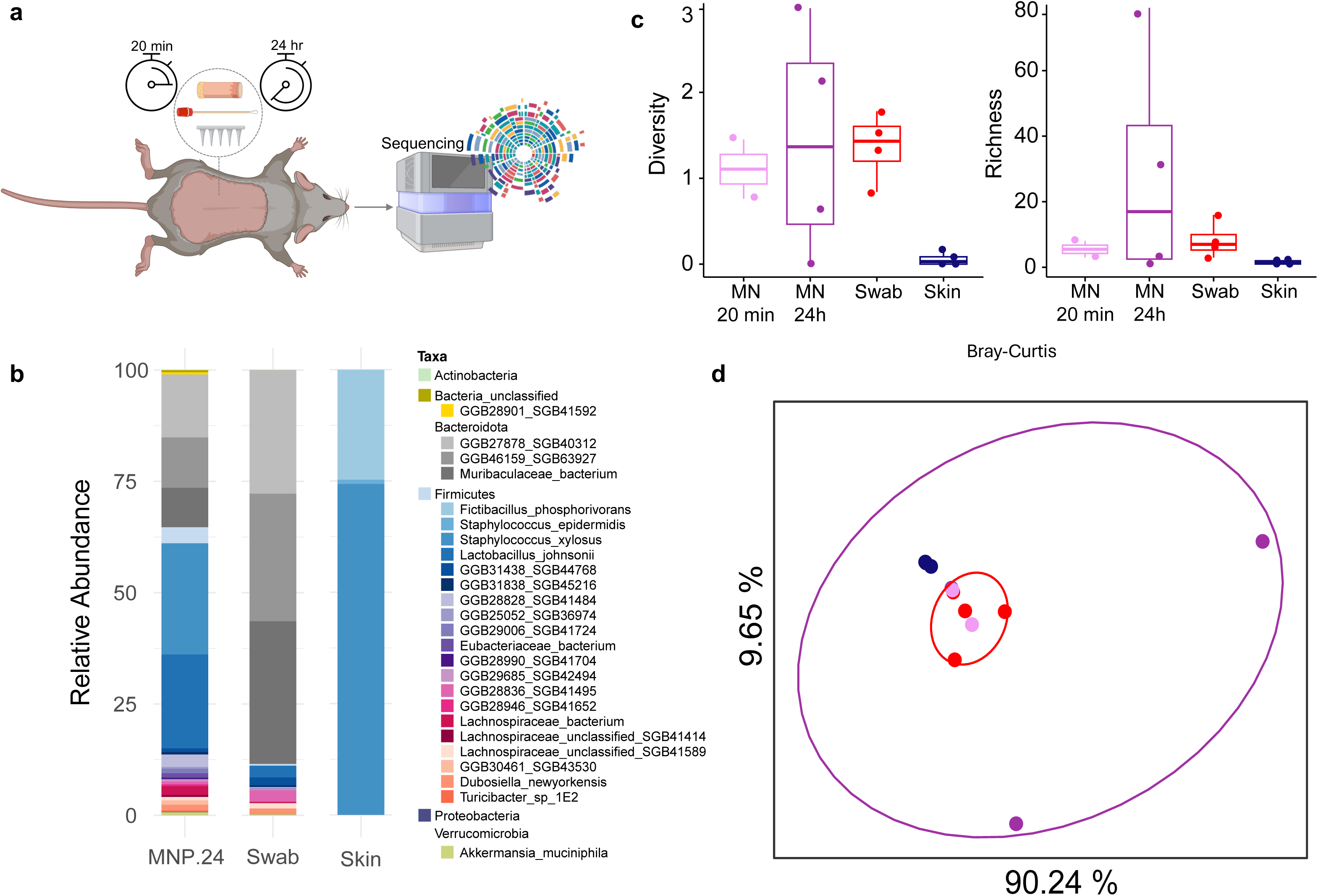
Microneedle sampling recapitulates murine skin microbial community structure. **a**, Schematic view showing the murine skin microbiome sampling using the MN patches, swabs, and skin biopsies. **b**, Relative abundance plots of the 25 abundant taxa are shown aggregated for all 24-hour MN patches, skin swab and skin tissue. Remaining species were grouped by their respective (**c**) phylum alpha diversity and species richness. **d**, PCOA clustering based on Bray-Curtis dissimilarity index (beta diversity) showing the samples collected from skin swabs, 20 min patches, 24-hour patches and the skin tissue. n=5 animals per group were used for all panels.

### Human MN sampling preserves inter-individual microbial signatures

We evaluated the feasibility, tolerability, and microbiome sampling performance of MN patches in a diverse human cohort. MN patches were applied for 20 minutes or 24 hours to the arm and upper back/neck region of eight healthy individuals spanning both sexes, multiple racial backgrounds, and a broad age range (**Fig. 3a,b; Supplementary Fig. S3**). Matched swab samples were collected from the same anatomical locations for comparison, followed by 16S rRNA sequencing. Participant-reported tolerability was assessed immediately following patch application. Across all subjects, MN insertion was well tolerated, with minimal pain scores and no reports of significant irritation or discomfort (**Fig. 3c**). No subject reported adverse effects during either 20-minute or 24-hour wear periods. Macroscopic examination of application sites before insertion, during wear, and after removal revealed no signs of aberrant erythema, edema, or sustained inflammation (**Fig. 3d**). Mild transient impressions at the insertion sites resolved rapidly without visible tissue disruption. These findings demonstrate that MN-based sampling was well tolerated in this study, supporting its suitability for longitudinal human studies. Sequencing depth was comparable across MN and swab samples, with read counts exceeding negative controls by several orders of magnitude (**Supplementary Fig. S4a**), suggesting sufficient microbial recovery, robust library complexity, and minimal background contamination. Air and extraction controls exhibited distinct compositional profiles relative to biological samples (**Supplementary Fig. S4c**), supporting data integrity. As previously, reads corresponding to Flavobacterium, identified in reagent controls, were excluded from downstream analyses (**Supplementary Fig. S4b).** Compositional analysis revealed that Actinobacteria, Firmicutes, and Proteobacteria were the dominant phyla across all sampling modalities, consistent with established human skin microbiome datasets. *Staphylococcus* was the most abundant genus across anatomical sites (**Fig. 3e; Supplementary Fig. S5a**), irrespective of sampling technique. Shannon diversity indices were comparable between MN and swab samples (**Fig. 3f)**, although swabs exhibited slightly higher median richness. Beta diversity analyses demonstrated overlap between MN-derived samples and to a degree, swab-derived communities within the same anatomical site (**Fig. 3g**), indicating that sampling modality imposed relatively limited variation on overall microbiome structure. Notably, MN samples clustered more tightly than swab samples, suggesting a more homogeneous recovery across individuals (**Fig. 3g**). Arm and back samples also showed a degree of site-dependent segregation, particularly in the 24-hour MN condition, consistent with preservation of some anatomical differences in the human skin microbiome (**Supplementary Fig. S4d**). Relative abundance profiles across subjects further confirmed that MN samples closely recapitulated swab-derived phylum distributions while maintaining patient-specific microbial signatures (**Supplementary Fig. S5b**). Together, these findings show that minimally invasive MN sampling largely recapitulates microbiome community structures captured by conventional skin swabbing, with inter-individual and site-specific differences exceeding modality-dependent effects. Combined with good tolerability and rigorous contamination control, these data support MN-based multimodal sampling as a robust platform for longitudinal human microbiome and immune profiling.

**Figure 3.**
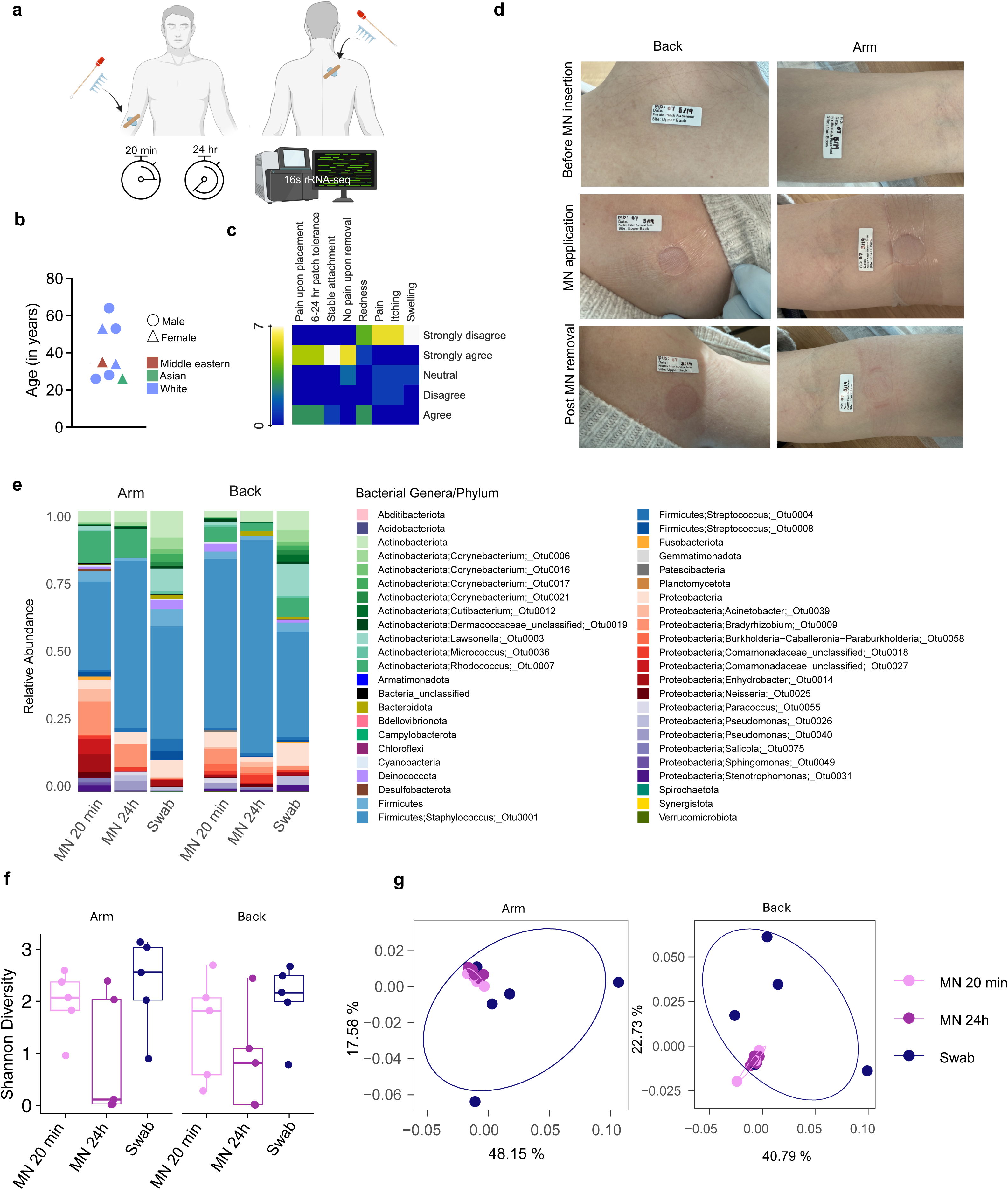
Microneedle sampling preserves inter-individual microbiome signatures. **a**, Schematic view showing the human skin microbiome sampling using the MN patches and skin swabs. **b**, Human subject demographics showing the age, gender and ethnicity. **c**, Tolerability of MN sampling as assessed on the cohort of volunteers (n=8). **d**, Representative photographs of pre- and post-MN patch application on the arm (dry, left) and back (sebaceous, right) of human volunteers. **e**, Community analysis of microbes captured with MN patches and swabs. Abundance plots of study participant samples at phylum level. **f**, Shannon diversity for patient samples (downsampled to min. patient sample read depth of 906 reads). **g**, PCOA (based on Bray-Curtis dissimilarity index) showing the samples collected from arms and the backs of individuals.

### Pathogen colonization induces progressive immune escalation captured longitudinally by MN sampling

Having established microbial sampling capability, we leveraged the multimodal platform to interrogate longitudinal immune responses during microbial dysbiosis. C57BL/6J mice were topically colonized with methicillin-resistant *S. aureus* for 1 or 3 days to establish an epicutaneous infection, followed by MN-based immune cell sampling and flow cytometric phenotyping (**Fig. 4a, Supplementary Fig. S6a**). Bacterial burden quantified via swabs and MN patches was comparable at day 3, whereas day 1 exhibited greater variability, likely reflecting variable colonization and superficial bacterial distribution prior to establishment of infection (**Fig. 4b**). Flow cytometry revealed time-dependent immune escalation within the colonized skin. Total CD45⁺ leukocytes increased significantly following colonization, accompanied by progressive expansion of CD11b⁺ myeloid cells, Ly6G⁺ neutrophils, F4/80^high^ macrophages, Ly6C⁺ inflammatory monocytes, and CD3⁺ T cells (**Fig. 4c,d**). While multiple subsets increased relative to controls at both timepoints, macrophages, neutrophils, and T cells were significantly elevated at day 3 compared to day 1, indicating sustained inflammatory amplification. Flow cytometry-based isolation of the same immune subsets from matched skin tissue digests demonstrated similar kinetics and magnitude of immune expansion (**Supplementary Fig. S6b**), confirming that MN sampling faithfully recapitulates cutaneous immune dynamics. In contrast, peripheral blood immune populations did not exhibit comparable magnitude or temporal escalation, with several subsets remaining relatively stable across timepoints (**Supplementary Fig. S6b**). These findings indicate that the observed immune amplification is tissue-restricted rather than driven by systemic leukocytosis, validating the spatial specificity of MN-based sampling.

**Figure 4.**
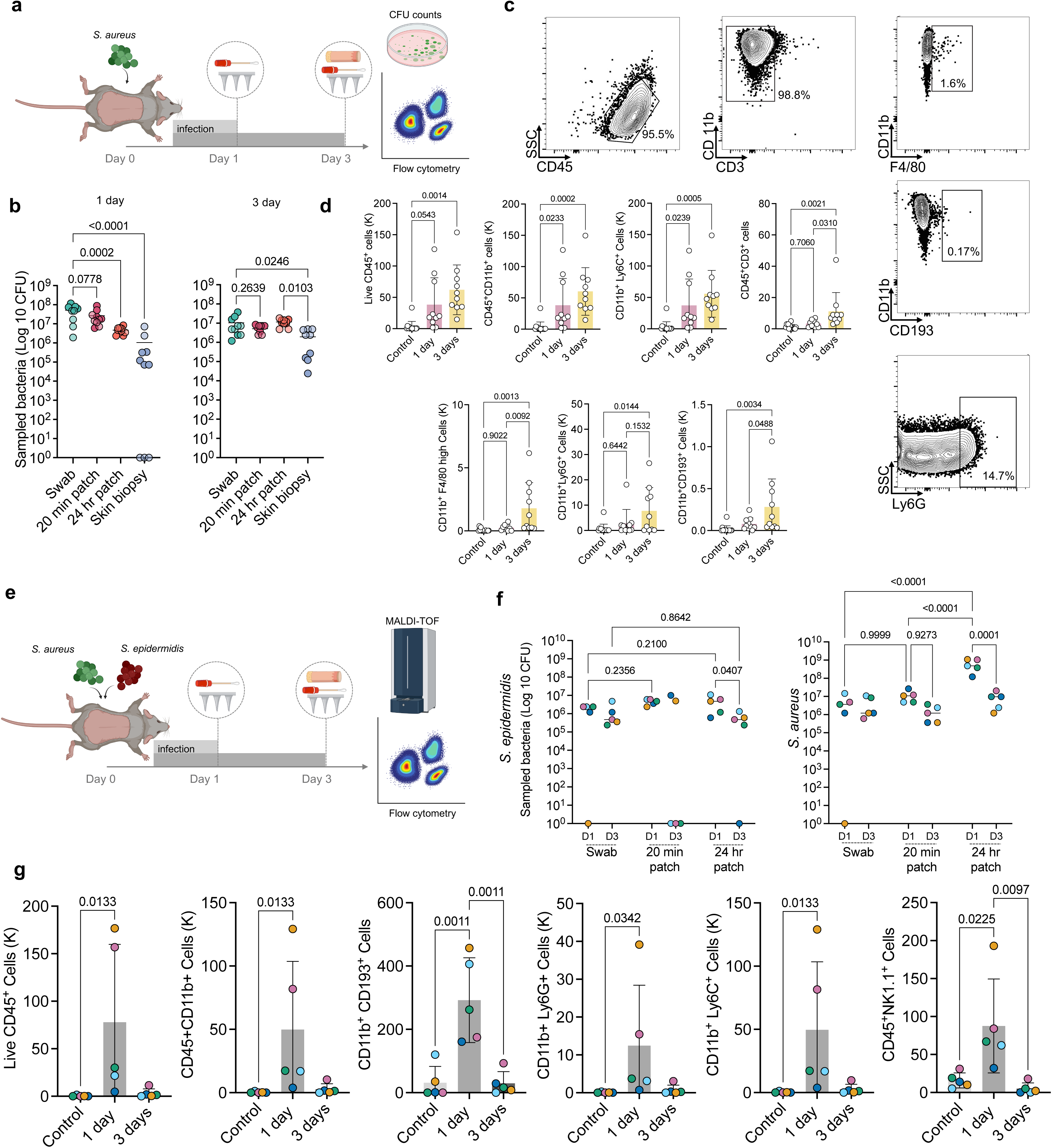
Longitudinal microneedle sampling reveals distinct immune trajectories during commensal and pathogen colonization. **a**, Schematic representation of the cutaneous *S. aureus* infection model and temporal sampling of bacteria and immune cells from mice. **b**, Comparison of *S. aureus* burden captured by different sampling modalities (Skin swabs, MN patches: 20 min and 24 hour and skin digest) from 1-day (left) and 3-day (right) infection (dots are color-coded by experimental group; lighter and darker shades within each color denote data collected from two independent experiments). **c**, Representative flow cytometry plots showing the skin-infiltrating myeloid cells, T cells, macrophages, eosinophils and neutrophils. **d**, Enumeration of recovered live leucocytes, myeloid cells, myeloid derived suppressor cells, T cells, macrophages, neutrophils and eosinophils from control group, 1-day and 3 days post *S. aureus* infection (n=10 animals per group). **e**, Schematic representation of the cutaneous dual bacterial colonization model using *S. epidermidis* and *S. aureus*. **f**, CFU counts of *S. epidermidis* and *S. aureus* temporally sampled by the MN patches from the mice counted and differentiated using MALDI-TOF. **g**, Enumeration of recovered live leucocytes, myeloid cells, eosinophils, neutrophils, myeloid derived suppressor cells and NK cells from control group, 1-day and 3 days post dual colonization of *S. epidermidis* and *S. aureus* (*n*=5 animals per group). (panel f and g: each colored dot represents an individual mouse, with bacterial burden and immune cell measurements derived from the same MN patch). Data shown are mean±SEM. P values were determined using one-way ANOVA (b,d,f) and two-way ANOVA (e) followed by Tukey’s multiple comparisons test. Panel a and e elements were created using BioRender.

### Commensal co-colonization reshapes pathogen-induced immune dynamics

To investigate how commensal bacteria might influence pathogen-driven inflammation in skin, mice were co-colonized with equal CFUs of *S. epidermidis* and *S. aureus* and monitored longitudinally (**Fig. 4e**). We quantified bacterial burden recovered by different sampling modalities during dual colonization with *S. epidermidis* and *S. aureus* at day 1 and day 3 post-inoculation (**Fig. 4f**). At day 1, 24-hour MN patches recovered higher *S. aureus* counts than 20-minute patches and swabs by 50 and 100 folds, respectively (**Fig. 4f**, right), consistent with the murine microbiome experiments. By day 3, the 24-hour MN patches revealed a marked reduction in *S. aureus* burden relative to day 1, with CFUs decreasing by nearly 150-fold (**Fig. 4f**), in contrast to the monocolonization model, in which *S. aureus* burden remained stable over time (**Fig. 4b**). Although bacterial counts tended to be higher with 24-hour patches, the difference between 20-minute and overnight MN sampling was not statistically significant at day 3.

Co-colonization induced a pronounced early immune surge at day 1, characterized by increased CD11b myeloid cells, Ly6G⁺ neutrophils, NK1.1⁺ cells, Ly6C⁺ monocytes, and myeloid-derived suppressor-like populations. Strikingly, these populations declined by day 3, in parallel with the reduction in *S. aureus* burden, in contrast to the progressive escalation observed during *S. aureus* monocolonization (**Fig. 4f**). Parallel immunophenotyping of matched skin tissue digests reproduced this transient kinetic pattern observed with MN sampling, although some immune populations remained elevated at day 3 in the tissue digest dataset (**Supplementary Fig. S6c**). This partial divergence likely reflects differences in the tissue compartments sampled by MN patches versus full skin digestion, while preserving the overall conclusion that co-colonization reshapes local immune dynamics over time. Peripheral blood, however, showed comparatively muted or non-parallel changes across the same timepoints (**Supplementary Fig. S6c**), reinforcing that the early surge and subsequent attenuation are localized tissue phenomena rather than systemic immune fluctuations. Notably, tracking bacterial and immune readouts within individual animals showed that mice with the highest residual *S. aureus* burden at day 3 also exhibited elevated total leukocyte, myeloid, and lymphoid responses by both MN patch and skin biopsy analyses (**Fig. 4f,g**; **Supplementary Fig. S6c**), further linking local bacterial persistence to the magnitude of tissue inflammation. This pattern is consistent with prior reports showing that *S. epidermidis* induces skin-adapted immune cell programs that enhance barrier immunity and limit pathogen invasion^11^. This transient activation pattern suggests that commensal presence modulates pathogen-induced immune responses, potentially through immune education, microbial competition, or altered inflammatory signaling. By enabling repeated sampling from the same anatomical site, MN-based multimodal profiling captures these dynamic immune-microbiome interactions *in vivo*, revealing tissue-restricted immune calibration that would be difficult to resolve using conventional terminal sampling approaches.

## Discussion

Here, we establish minimally invasive MN sampling as a platform for repeated, spatially defined profiling of microbiota and infiltrating immune cells from intact murine and human skin, and leverage this approach to uncover distinct temporal immune trajectories during pathogen monocolonization and commensal-pathogen co-colonization. Comparison with conventional sampling methods, such as skin swab and biopsy, revealed similar or superior results with MN patches in both preclinical animal models and in human studies. Our data reveal that co-colonization with *S. epidermidis*, a common member of the human skin microbiota, dynamically recalibrates pathogen-induced inflammatory escalation rather than simply reducing bacterial burden, highlighting how microbial context shapes inflammatory kinetics within barrier tissue.

Recent MN technologies have largely been developed for sampling soluble biomarkers from ISF, with some incorporating integrated biosensors or material-responsive elements to enable on-patch chemical detection^19–22^. A subset of transepidermal microprojection arrays has been applied to microbiome sampling; however, these approaches were focused on transepidermal fungal species detection or material-triggered analytical readouts, without resolving concurrent host immune dynamics^23,24^. Moreover, many existing systems are optimized for single-modality sampling and do not enable simultaneous recovery of immune cells and microbial DNA from the same cutaneous microenvironment^19,25^. In contrast, our platform integrates broad microbial capture with *in situ* immune cell profiling in a minimally invasive, FDA-compatible design, enabling longitudinal, site-specific interrogation of commensal-pathogen-host interactions in living tissue. This multimodal capability advances MN technology from descriptive or analyte-restricted sampling toward temporally resolved systems-level analysis of barrier immunity.

To optimize recovery efficiency, we systematically screened a panel of alginate hydrogel coatings with distinct physicochemical properties. We identified SLG20, characterized by a higher G/M ratio and lower molecular weight, as providing superior recovery performance. The high G-content of SLG20 promotes formation of a stiffer yet more porous “egg-box” network through Ca^2+^-mediated crosslinking of G-blocks, while its lower molecular weight reduces chain entanglement and increases effective pore size^26–28^. Together, these structural features enhance hydrogel swelling, fluid exchange, and cellular ingress, consistent with published findings demonstrating improved cell mobility and nutrient transport in high G-content, low-molecular-weight alginate matrices^29^. Notably, this same SLG20 composition was shown to exhibit superior immune cell sampling properties *in vivo*^30^, further supporting the concept that hydrogel microarchitecture is a key determinant of biological capture efficiency.

Our murine and human data together establish that this MN platform can capture the predominant skin microbiota captured by existing methods while recapitulating overall community composition. In mice, extended versus short 20 min applications MN application improved recovery from dorsal skin and yielded communities with substantial overlap with those detected by swabs and biopsies. In human subjects, MN sampling similarly preserved the dominant taxonomic patterns and inter-individual signatures observed with swab-based profiling, supporting its utility for minimally invasive longitudinal microbiome analysis across species. Consistent with these findings, *in vitro* structural validation, depth-dependent sampling experiments, and viability analyses indicate that microbial recovery suggests a controlled tissue penetration rather than passive surface adsorption or hydrogel-associated toxicity. An important next step will be to define how tissue depth and local skin structure influence MN-based microbial recovery *in vivo*. Future studies integrating controlled MN geometries with histology, depth-resolved imaging, or spatially registered microbial measurements could determine whether differences in sampling duration across species partly reflect differences in epidermal thickness and skin organization.

In murine metagenomic datasets, several sequences could not be confidently assigned at strain level, reflecting current limitations in reference databases for mouse skin microbiota rather than platform performance^31–33^. Continued expansion and curation of mouse-associated microbial databases will improve taxonomic resolution. In a diverse human cohort, MN application was well tolerated, with minimal discomfort and no sustained erythema, edema, or tissue disruption. Microbiome profiling revealed that *Actinobacteria*, *Firmicutes*, and *Proteobacteria* remained dominant phyla and that MN-derived communities preserved individual- and site-specific signatures comparable to swabs, with beta diversity driven primarily by anatomical site and subject identity rather than sampling modality. The tighter clustering of MN samples further suggests a more consistent microbiome recovery, although the lower diversity recovered also raises the possiblity that MN sampling captures a narrower breadth of microbes. 24-hour MN sampling better preserved site-dependent segregation between arm and back samples over short samplings. Although validation in larger cohorts and across additional anatomical sites is needed, these findings support the promise for MN sampling for longitudinal human microbiome studies. To our knowledge, this represents the first multi-individual study using sampling MN patches for native human skin microbiome interrogation, building on prior devices that monitored immune cells or ISF but did not capture microbiota. Although both short and extended wear periods yielded recoverable microbial DNA, shorter application times may be preferable in many clinical settings to minimize burden and improve compliance. Importantly, we demonstrate that a 20 minute wear time is sufficient for both microbiome capture and ISF sampling of cytokines and chemokines^30^. The robust performance of 20-minute MN sampling in human skin suggests that short wear times are sufficient to recover stable and subject-specific microbial signatures for human skin, at least. The distinct duration dependence observed between human and murine skin may reflect species-dependent differences in skin structure, microbial density, and surface organization, which will require direct testing in future studies.

We demonstrated that MN arrays can recover viable bacterial communities alongside infiltrating immune cells from the same location *in vivo*. We leveraged this multimodal platform to interrogate how pathogen-only versus dual colonization with *S. epidermidis* and *S. aureus* shape local immune trajectories over time. We observed divergent immune programs during colonization. In the *S. aureus* model, serial sampling revealed progressive amplification of CD45⁺ leukocytes, with expansion of CD11b⁺ myeloid cells, Ly6G⁺ neutrophils, F4/80^high^ macrophages, Ly6C⁺ inflammatory monocytes, and CD3⁺ T cells between day 1 and day 3, consistent with sustained inflammatory escalation in the skin but not in peripheral blood. These kinetics are consistent with prior reports that *S. aureus* infection elicits robust myeloid and T cell responses in the skin^34,35^, but here we resolve how these responses accumulate over time within the same microenvironment. By directly aligning microbial burden with evolving immune composition at fixed cutaneous loci, our data move beyond inference to a temporally linked view of local infection dynamics. By contrast, co-colonization with *S. epidermidis* reshaped these dynamics in a manner consistent with, but not reducible to, classical commensal protection models. Rather than simply decreasing *S. aureus* burden, the presence of *S. epidermidis* induced a pronounced early surge of myeloid and lymphoid populations at day 1, followed by contraction by day 3 despite persistence of both organisms. This transient peak suggests that commensals may modulate the temporal architecture of inflammation, promoting rapid activation and subsequent attenuation, rather than uniformly dampening immune responses. Prior studies have shown that *S. epidermidis* colonization can induce tissue-resident T cells, enhance keratinocyte antimicrobial defenses^11,36^, and constrain *S. aureus* through strain-specific antimicrobial factors^37^, providing potential mechanistic frameworks for such temporal modulation. In addition, interspecies signaling or quorum interference may attenuate *S. aureus* virulence programs during co-colonization, thereby reducing inflammatory output without eliminating bacterial persistence. A key limitation is the absence of an *S. epidermidis*-only condition, which precludes full separation of co-colonization effects from those induced by *S. epidermidis* alone. Although we did not directly quantify cytokine pathways, antimicrobial peptides, or bacteriocin activity here, our observations are compatible with models in which commensal-primed cellular and antimicrobial circuits accelerate early pathogen control, allowing earlier attenuation of cellular infiltration at the same site. These data thus raise testable hypotheses that commensal-induced T cell programs, barrier-associated antibodies, and microbial competition collectively help set an upper bound on inflammatory escalation during co-colonization. [I think you need to mention a caveat though, that you don’t have the S. epidermidis only data which I think is important)

The capacity to co-sample immune cells and microbiota directly addresses key limitations of conventional swabs and biopsies. Conventional skin microbiome sampling methods, including swabbing, tape-stripping, and biopsy, present significant limitations. Swabs provide practical assessment of surface microbial composition but do not capture infiltrating immune populations or soluble mediators within the epidermis, and they predominantly sample only the superficial bacterial populations with limited depth control and high variability in collection efficiency, preferentially sampling superficial strata^38–40^. Biopsies allow deep immune and molecular profiling but are invasive, disrupt the local niche, and are poorly suited to repeated sampling, limiting their use in longitudinal studies and in healthy populations^38,41^. Other minimally invasive methods, such as tape stripping and suction blistering, can recover superficial cells, soluble mediators, and nucleic acids, but provide limited depth control, often perturb barrier integrity, and typically do not capture viable bacteria and immune cells from the same microenvironment^39,42^. By contrast, our MN platform fills a distinct niche by co-sampling viable immune cells and native microbial communities, with sufficient yield for culture, metagenomics, and multiparameter cytometry, while preserving the tissue architecture for repeated interrogation. This capability is particularly relevant in settings such as atopic dermatitis flares, vaccine-site responses, engineered commensal colonization, and age-associated changes in barrier immunity, where shifts in microbial composition and local immune state are tightly coupled. Positioning the platform within this broader landscape emphasizes that it complements, rather than replaces, existing tools, and uniquely enables kinetic, site-matched host-microbe studies.

Larger longitudinal studies across diverse anatomical sites and disease contexts, including aging, autoimmune skin disorders, infectious and wound-healing settings, vaccine responses, and microbial transplantation paradigms, will be necessary to define population-level variability and mechanistic generalizability. Future iterations integrating shotgun and strain-resolved metagenomics, isolate sequencing, locus-specific qPCR, high-dimensional cytometry, proteomics, and single-cell or spatial transcriptomics will enable alignment of microbial genomic features with cellular, humoral, and barrier-associated trajectories. In parallel, incorporating quantitative barrier measurements, such as transepidermal water loss, corneometry, and spatial profiling of keratinocyte differentiation and stress pathways, will allow direct testing of how transient immune activation and contraction couple to barrier disruption or repair in inflammatory disease.

The ability to longitudinally co-sample microbiota and immune cells from defined cutaneous sites creates immediate opportunities for mechanistic experimentation and clinical translation. Applying MN-based sampling to controlled *S. epidermidis* colonization in wild-type and immune-deficient mice, with concurrent measurement of local antibody and cytokine concentrations, would directly test how skin-autonomous humoral and cellular programs shape the transient activation and contraction we observe ^11,43^. In human disease, therapeutic introduction of protective commensals could be monitored in real time to determine whether successful colonization shifts immune trajectories from escalating to self-limited inflammation. Similarly, at vaccine sites, MN patches could track how resident microbiota influence tissue-resident T cell recruitment, cytokine milieus, and local antibody production, informing rational adjuvant or commensal co-therapies. The platform could also be deployed in trials of biologics, JAK inhibitors, topical small molecules, or wound-healing interventions to map how targeted immune modulation reshapes microbial communities and cellular recruitment within lesional and non-lesional skin. More broadly, integrating MN-based longitudinal readouts into interventional studies would enable mechanism-guided patient stratification and dynamic response monitoring, accelerating translation of microbiome and immune insights into personalized, tissue-informed therapies.

Collectively, this work reframes MN sampling not simply as a technological advance but as an enabling strategy to study temporal host-microbe dynamics within intact tissue. By revealing distinct immune trajectories during pathogen alone versus commensal-pathogen co-colonization, our findings underscore that cutaneous immunity operates as a dynamic, context-dependent system rather than a static response to microbial presence. Approaches that permit minimally perturbative, repeated sampling of the same microenvironment, such as the MN patches described here, will be essential for building mechanistic models of how barrier tissues maintain symbiosis, transition to inflammation, and restore homeostasis, and for translating those models into rational microbiome- and immunity-based interventions in humans. Although the present study focuses on skin, the adaptable architecture of these patches positions them for future application to other barrier and mucosal sites, including the oral and vaginal microenvironments, where immune-microbiome interactions similarly shape tissue health and disease.

## Methods

### Fabrication and characterization of MN patches

Polydimethylsiloxane (PDMS) templates (Sylgard 184; Dow-Corning) were produced using laser micromachining technology (Blueacre Technology, Ireland). Poly(L-lactide) (PLLA; RESOMER L 207 S, Evonik Industries AG) was subsequently melted and cast onto the molds under vacuum conditions (−20 mmHg) at 200 °C for 120 minutes to obtain solid MN arrays. The resulting patches were subjected to oxygen plasma treatment (Diener Electronic, Germany) for 2 minutes at 0.5 mbar to enhance surface hydrophilicity. A 0.58% (w/v) sodium alginate solution (PRONOVA SLG20, SLM20, SLG100, or SLM100; Novamatrix, IFF) supplemented with 4.6% (v/v) sucrose (Teknova, S00572PK) was then evenly dispensed over each patch and air-dried at ambient temperature (25 °C) for a minimum of 4 hours. Cross-linking was performed by applying a 20 mM CaCl_2_ solution (Sigma, 21115) onto the alginate-coated surfaces, followed by overnight drying in a sterile laminar flow hood. The geometry and morphology of the MN arrays were examined using a Zeiss Sigma 360 scanning electron microscope. For samples used *in vivo*, tissues containing the MNs were fixed in 4% paraformaldehyde (Electron Microscopy Sciences, 157-4) and 2.5% glutaraldehyde (Sigma, G7776) for 2 hours, post-fixed with 0.5% osmium tetroxide (Electron Microscopy Sciences, 19152) for 1 hour and dehydrated through a graded ethanol series. Specimens were subsequently dried overnight prior to imaging.

### Bacterial cultures and biotyping

*Staphylococcus aureus* USA300 LAC (strain USA300-0114; BEI Resources, NR-46070) and *Staphylococcus epidermidis (ATCC12228; ATCC)* was streaked on blood agar and incubated overnight at 37 °C. A single colony was used to inoculate tryptic soy broth (TSB; Fisher Scientific, DF0370-17-3) and cultured overnight at 37 °C with orbital agitation (200 rpm). The relationship between optical density (OD_600_) and viable cell count was established by enumerating colony-forming units (CFU) per milliliter using a hemocytometer (Fisher Scientific, 22-600-107) in triplicate. One OD_600_ unit corresponded to approximately 2×10^8^ CFU/mL. For experimental use, overnight cultures were diluted 1:200 into fresh TSB and grown to mid-logarithmic phase (OD_600_ ≈ 1) under the same conditions. Bacteria were harvested by centrifugation, washed once with sterile phosphate-buffered saline (PBS), and pelleted again. The final bacterial suspension was resuspended and adjusted in PBS to a concentration of 10^8^ CFU per 150 µL.

### Bacterial viability assays

An overnight culture of GFP-expressing *S. epidermidis* was mixed with alginate (SLG20) and drop-cast into a tube containing 20 mM CaCl_2_ solution to induce gelation. The resulting contents were passed through a cell strainer, and the alginate beads were subsequently decrosslinked using 20 U alginate lyase. Bacterial cells were collected by centrifugation, resuspended, serially diluted, and plated for CFU determination. An equivalent volume of the overnight culture without alginate served as a positive control. For imaging assays, 20 µL suspension of alginate-bacteria was placed at the center of a 96-well culture plate, and the resulting dome was crosslinked by adding 20 mM CaCl_2_ to the wells. After crosslinking, the excess CaCl_2_ solution was removed, and the wells were rinsed twice with PBS before imaging GFP fluorescence. Following imaging, TSB medium was added to each well, and the plate was incubated overnight at 37°C. The next day, the medium was removed, the wells were washed with PBS, and fluorescence images were acquired to assess bacterial growth. Mean GFP fluorescence intensity was quantified using ImageJ software.

### *In vitro* bacterial sampling

Bacterial suspensions (∼1×10^8^ CFU/mL) were prepared from fresh overnight cultures. One milliliter of each suspension was plated onto blood agar and evenly spread using an L-shaped spreader. Low-melting-point agarose (Lonza Bioscience, Cat. No.50101) was prepared at 2% w/v in sterile PBS by heating to 50□°C until fully dissolved. The solution was then cooled to 40□°C, and 10□mL was poured over the bacterial lawn to fully cover the plate and allowed to solidify. MN patches were subsequently applied onto the agarose layer, ensuring needle penetration through the agarose to contact the bacterial lawn. After 20□minutes, the patches were carefully removed and processed for CFU enumeration. To assess the bacterial sampling capacity of MN patches with different penetration depths, MNs of varying lengths (600 and 1000 µm), were tested. The agarose volume was adjusted to form a 700 µm-thick layer, and MNs of each length were applied to this layer. Bacterial CFU enumeration was then performed as described above.

### Mice

Female *C57BL/6J* (JAX 000664) mice were obtained from The Jackson Laboratory (Bar Harbor, ME, USA). Mice were at the age of 6-8 weeks, at the onset of experimentation. Animals were housed in the University of Connecticut Health Center (UCHC) animal facility under specific pathogen-free conditions, with controlled temperature and humidity and a 12-hour light-dark cycle. Standard chow and water were provided *ad libitum*, and up to five mice were co-housed per cage. All animal procedures were conducted in compliance with protocols approved by the UCHC Institutional Animal Care and Use Committee (IACUC) and adhered to the National Institutes of Health guidelines for the care and use of laboratory animals. Mice were euthanized at study endpoints using CO_2_ asphyxiation in accordance with institutional ethical standards.

### Bacterial colonization model in mice

Epicutaneous colonization with *Staphylococcus aureus* and *S. epidermidis* was established following previously described approaches^44^ with minor modifications. Briefly, dorsal hair of mice was removed using electric clippers and a depilatory agent (Veet; Reckitt) at least 48 hours prior to infection to allow for skin recovery. On day 0, animals were anesthetized as detailed above. A sterile, non-stick gauze pad (1.5 × 1.5 cm; CVS Health) was saturated with 150 µL of *S. aureus* suspension (1×10^8^ CFU in PBS) and placed on the exposed dorsal skin. The gauze was covered with a semipermeable bio-occlusive dressing (Tegaderm; 3M) and secured with two layers of adhesive bandage to maintain contact. Mice were inspected daily for signs of distress or dressing displacement, and outer bandages were replaced as needed. After 72 hours, under anesthesia, dressings and gauze pads were carefully removed for subsequent analyses.

### Murine bacterial and Immune cell sampling

Microneedle (MN) patches were applied to the shaved dorsal skin of anesthetized mice. Each patch was gently pressed onto the skin for approximately 10 seconds to ensure contact and then secured using a Tegaderm (3M) and adhesive bandages. For immune cell and bacterial collection, MN patches were applied for either 20 minutes or overnight. For overnight applications, patches were additionally reinforced with electrical tape to prevent removal by cohoused animals. Mice were observed continuously after application and monitored daily thereafter for any signs of distress. To retrieve the patches, animals were briefly re-anesthetized, and the coverings were carefully removed. Recovered patches were placed in 12-well plates containing 20 U/ml alginate lyase solution supplemented with 10% fetal bovine serum (FBS) for immune cell recovery. Plates were agitated on a shaker for 20 minutes to dissociate alginate crosslinks. Following agitation, patches were gently held against the well wall with sterile forceps and rinsed thoroughly with the same elution buffer. The well contents containing the sampled bacteria and immune cells were transferred into U-bottom 96-well plates for subsequent CFU measurements and flow cytometry analysis. The 96 well plate containing MN eluent was centrifuged at 1500 rpm for 10 minutes at 4°C. The supernatant was carefully transferred to a new well plate. The pellet containing immune cells were further stained for flow cytometry analysis.

### Bacterial CFU count assessment

The well plate containing the supernatant from the MN eluent was centrifuged at 15,000 rpm to pellet bacterial cells. The supernatant was removed, and the pellet was resuspended in 100 µL of tryptic soy broth (TSB). Tenfold serial dilutions were prepared, and 10 µL from each dilution was spot plated onto tryptic soy agar (TSA) plates. Plates were incubated at 37 °C overnight, and colony-forming units (CFUs) were enumerated and back-calculated to determine the bacterial load per sample.

### PBMC Isolation

Peripheral blood was collected before and after *S. aureus* colonization for flow cytometric analysis. Under anesthesia, mouse blood was obtained via retro-orbital puncture using a heparinized capillary tube (Drummond, 21-176-6) and transferred into K₂EDTA-coated vacutainer tubes (BD Biosciences, 367856). Tubes were gently inverted to ensure anticoagulant mixing and maintained at room temperature until red blood cell (RBC) lysis. Whole blood was diluted with ten volumes of 1× RBC lysis buffer and incubated at room temperature for 4-5 minutes with gentle agitation. The lysis reaction was quenched by adding three volumes of 1× PBS, followed by centrifugation at 300-400 *g* for 5 minutes at 2-8 °C. The lysis step was repeated twice to ensure complete erythrocyte removal. The resulting cell pellet was resuspended in FACS buffer and transferred to U-bottom 96-well plates for downstream flow cytometric analysis.

### Skin tissue dissociation and processing

Excised mouce skin samples were weighed prior to processing. Subcutaneous adipose tissue was carefully removed from approximately 0.5 × 0.5 cm sections, which were then finely minced using sterile scissors. The tissue fragments were transferred into 5 mL centrifuge tubes containing 1 mL of enzymatic digestion buffer composed of 2 mg/mL collagenase XI (Sigma, C9407-1G), 0.5 mg/mL hyaluronidase (Sigma, H3506-1G), and 0.1 mg/mL DNase I (Sigma, DN25-10MG) prepared in RPMI medium supplemented with HEPES and penicillin– streptomycin. An additional 2 mL of the same digestion mixture was added, and samples were further minced to enhance enzymatic exposure. Tubes were incubated in a shaking incubator at 37 °C for 45 minutes at 255 rpm. Following digestion, the resulting cell suspension was filtered through a 100 µm cell strainer into a 50 mL conical tube containing RPMI with 10% fetal bovine serum (FBS) and penicillin-streptomycin to neutralize enzyme activity. The filtrate was centrifuged at 1,500 rpm for 5 minutes at 37 °C. The resulting cell pellet was resuspended in cell sorting buffer and transferred to U-bottom 96-well plates for flow cytometric analysis. Final cell counts were normalized to 100 mg of processed skin tissue.

### Flow cytometry analysis

Cells isolated from peripheral blood, MN patches, and skin tissue were transferred into 96-well U-bottom plates and centrifuged at 1,200 rpm for 5 minutes to pellet the cells. Supernatants were aspirated, and pellets were resuspended in 100 µL of fixable Live/Dead Zombie NIR viability dye (BioLegend, Cat. 423106; 1:1,000 dilution in PBS). Samples were incubated for 15 minutes at room temperature in the dark to assess viability. Following incubation, cells were washed once with FACS buffer and centrifuged at 1,500 rpm for 5 minutes. The resulting pellets were resuspended in 100 µL of anti-Fc receptor blocking antibody (clone 2.4G2) and subsequently stained with fluorochrome-conjugated antibodies diluted in PBS containing 2% fetal calf serum. Surface staining was performed for 30 minutes on ice in the dark. After staining, cells were washed with FACS buffer, resuspended in 200 µL of buffer, and analyzed by flow cytometry. Data acquisition was performed using a BD Symphony A5 cytometer (BD Biosciences) with *FACSDiva* software (BD Biosciences), and data were analyzed using *FlowJo* (BD Biosciences). The following antibodies were used for surface staining: CD45 (BUV395, Cat. 564279, BD Biosciences), CD3ε (AF488, Cat. 100321, BioLegend), CD11b (BV785, Cat. 101243, BioLegend), NK1.1 (BV421, Cat. 108732, BioLegend), F4/80 (PE, Cat. 123110, BioLegend), CD193 (APC, Cat. 144512, BioLegend), CD11c (BUV661, Cat. 750482, BD Biosciences), Ly6C (RB705, Cat. 570264, BD Biosciences), MHC-II (BUV496, Cat. 750281, BD Biosciences), and Ly6G (BUV805, Cat. 741994, BD Biosciences).

### Human study design

8 healthy adult volunteers (4 female and 4 male) with ages ranging from 26 to 64 years old (M_age_ = 39.9, SD = 14.7) were recruited at the Jackson Laboratory for Genomic Medicine under the Institutional Review Board-approved protocol (IRB #2025-005-JGM). Race and ethnicity were self-reported. Of the eight participants, six identified as White, one as Asian, and one as Middle Eastern. For ethnicity, seven participants identified a Non-Hispanic or Latino, and one identified as Hispanic or Latino. Participants were enrolled following eligibility prescreening and informed consent. Participants were prescreened for eligibility based on inclusion and exclusion criteria, including absence of skin conditions (e.g., presence of inflammation or lesions), recent topical treatments on the skin sites to be swabbed, use of systemic steroids, antibiotics, or antifungals within one month of sampling, immunosuppression, or severe, potentially confounding illnesses (e.g., diabetes requiring insulin, severe autoimmune disease). Upon confirmation of eligibility and consent, participants were scheduled for sample collection visits and were provided with instructions for skin preparation, including avoidance of bathing, emollients, creams, and antimicrobial soaps on sampling sites for 24 hours prior to Visit 1. At Visit 1, participants completed a series of questionnaires administered in person. These forms included questions regarding participant demographics, medical history, recent medication use, and current skin condition. Following questionnaire administration, participants underwent microbiome sampling at two anatomical sites: the inner elbow and the back of the neck.

### Human microneedle sampling

For each site 2 sets of four MN patches were firmly pressed onto the skin for 30 seconds and secured using Tegaderm and a bandage. Patches were removed at four time points: 20 minutes (Visit 1), 3-4 hours (Visit 2), 7-8 hours (Visit 3), and 24 hours (Visit 4). After removal, each patch was placed into a 12 well plate containing 20U/ml alginate lyase solution on ice. Photographs of the skin sites were taken before and after MN patch placement and removal to document skin condition. Upon completion of MN patch sampling, participants completed a questionnaire that involved rating a series of statements regarding patch wear experience and any skin reactions on a five-point Likert scale. An example item includes asking participants if “the patch was not painful upon placement”. Follow-up contact with the participants occurred within 1-2 business days after patch removal to assess for adverse events. Prior to MN patch application, each site was swabbed using a sterile swab dipped in PBS for microbiome analysis. Swabs were gently rubbed over a 4 cm^2^ area for 20-30 seconds and immediately placed into lysis buffer for processing. An environmental air swab was also collected as a negative control. At each site, up to 4 MN patches were applied to assess the microbiome sampling efficiency of the patches across varying time points. Two skin swabs were collected from each of the five healthy individuals included in the microbiome analysis cohort. A total of 30 patient samples plus paired “air swabs” as negative controls were 16S sequenced. One alginate and one non-alginate treated MN patches as well as 2 extraction negative controls were included.

### 16s rRNA gene sequencing

DNA was extracted from human study samples, including skin swabs, air swabs, and MN patch eluents, using the PureLink™ Genomic DNA Isolation Kit (Thermo Fisher Scientific, Cat. K182001) following the manufacturer’s instructions. Purified DNA samples were submitted to the UConn Microbial Analysis, Resources, and Services (MARS) facility for library preparation and 16S rRNA sequencing. 16S library preparation was carried out using barcoded primers targeting the V1-V3 region of the 16S rRNA gene (FWD 5’-AGAGTTTGATCCTGGCTCAG-3’, REV 5’-ATTACCGCGGCTGCTGG-3’). An in-house pipeline based on Mothur^45^ was used to quality screen, data cleaning, and classify the sequencing data. OTU_2 was the most abundant OTU identified in unused control MP only samples and recurring in the MN patches applied to skin. OTU_2 was classified as Flavobacterium and a contaminant from the alginate used to prepare the MN patches.OTU_2 was subtracted from the dataset. In addition, OTUs of the Halobacter taxon were removed from the dataset since they occurred in the negative controls (Halobacter). Sequencing depth across samples after contaminant removal is shown in Figure S4a. All patient samples had > 900 reads after contaminant removal with a median of 7011 and mean 35394. All taxonomic features with a mean relative abundance of <0.001% (denoise function, Arumugam et al.)^46^ across the dataset were removed to reduce potential false positives yielding 519 OTUs. No prevalence cutoff was applied due to the small sample size. Metadata is listed in Table S1.

### Whole genome sequencing data processing

We sequenced a total of 28 samples consisting of samples collected with MNs, swabs and skin biopsies as well as positive and negative controls. The metagenomes had a median of 10.9 million raw reads (for each R1 and R2 read) with a substantial mouse DNA admixture (median 77% of total reads). A median of 1.2 million reads remained after trimming, dehosting, and quality control. Raw reads were deduplicated with prinseq-lite-0.20.4 and then processed with Trimmomatic 0.39 (Bolger et al., 2014) to remove low-quality reads and Nextera adapter. Bowtie2 was used to map the reads to the mouse genome. Following, the quality-controlled and dehosted fastq files were processed with MetaPhlAn 4 (Blanco-Miguez et al., 2022). Metaphlan was able to classify 1.0 – 90.7 % of the reads in all but 8 samples in which no reads were classified. These samples were excluded from the analysis, 4 of these were negative controls and supposed to yield no data. Pooled assembly (without the mock positive control) was performed using Megahit. Contigs were then mapped to the nt database (NCBI) and the Kraken standard database using Kraken. abundance tables were used to investigate if the assembly improved the taxonomic resolution of the mouse skin microbiomes (data not shown).

### Analysis of Whole genome sequencing data

All species level data was extracted from the Metaphlan 4 output and species with a mean relative abundance of 0.001% (denoise function)^46^ across the dataset were included. Like for the human data, Flavobacterium (Figure S2b) was excluded from the analysis as it was deemed a contaminant from the MN patches. *Cutibacterium acnes* was abundant in few MN patches and removed as contaminant. The remaining proportions were scaled to 100% (prop.table, R function). The vegan package was used to calculate richness and Shannon diversity, and Bray-Curtis dissimilarity matrix^47^ (vegdist, vegan R package) with a pseudo count of 10^□7 and □log10 transformation was used for clustering of the data and then depicted as a principal coordinate Analysis (PCoA) plot. Ellipses were drawn using stat_ellipse (ggplot2 R package), showing the 95% confidence interval.

### Statistical analysis of sequencing data and visualization

Statistical analyses were performed using R (version 4.5.1; R Foundation for Statistical Computing) and RStudio (version 2025.09.0), as well as GraphPad Prism (GraphPad Software). For microbiome sequencing data, analysis and visualization were conducted using the R packages reshape2, vegan, ggplot2, tidyverse, ggpubr, dplyr, and plyr^16–20^. Corrected P values < 0.05 were considered statistically significant. For richness and Shannon diversity analyses, data were rarefied (rarefy function, vegan package) to the lowest read count among patient samples (906 reads). Comparisons between sample types were performed using the Wilcoxon test with false discovery rate (FDR) correction. For all other experimental datasets, statistical comparisons were performed using unpaired Student’s t-tests or one-way ANOVA followed by Tukey’s honestly significant difference (HSD) test for multiple comparisons. P values < 0.05 were considered statistically significant. Data are presented as mean ± SEM.

## Data availability

The main data supporting the results in this study are available within the paper and its Supplementary Information. The raw and analyzed datasets generated during the study are available for research purposes from the corresponding authors on reasonable request.

## Acknowledgements

We thank Rachael Norris for assistance with SEM imaging. We are grateful to Alan Ahlberg and Mindy Carpenter for their help with human studies and to Garrett Henry for support with 3D printing and MN patch fabrication. We also thank Michael Michaud for assistance with animal studies and Tiffany Prosio for help with flow cytometry. We appreciate the contributions of Kendra Mass to 16S rRNA sequencing and analysis, and Angela Elwell for assistance with metagenomic experiments. We thank Sara Cassidy and Mark Adams for editing and critical review of this manuscript. This work was supported by the National Institutes of Health (U01-AG084765-02 award to J.O. and S.J.) and the Pepper Center Scholar Award (P30AG067988, to S.J.). All schematic illustrations were created with BioRender.com

## Author contributions

A.K.D. and S.J. conceived and designed the experiments. A.K.D performed all the experiments. A.K.D. and S.Y.K. performed all *in vivo* experiments. A.Y.V. performed the analyses of murine and human microbiome sequencing data. J.O. supervised the microbiome sequencing studies and data analysis. A.K.D., S.G., and S.Y.K. conducted the human microbiome sampling studies. A.K.D. and S.J. wrote the manuscript with contributions from A.Y.V., S.G., and J.O. All authors reviewed and edited the manuscript. S.J. supervised the overall study.

