## Supplementary Information for "Longitudinal microneedle sampling resolves tissue-restricted immune-microbiome dynamics in skin"

**Table of contents**

**1. Supplementary Tables**

**2. Supplementary Figures**

### Supplementary Tables

#### Supplementary Table S1 | 16s rRNA gene sequencing metadata

| File | type.general | type | location | Patient | SampleID.short | SampleID |
|---|---|---|---|---|---|---|
| FASTQ_Generation_2025-06-28_18_53_06Z-836062246.2A20_L001-ds.295609fbc3f840cd9e38ac07e1e0f8f3.skinmicrobiome1_S1_L001 | MN | MN 20 min | Arm | P2 | 2A20 | 2A20.skinmicrobiome1 |
| FASTQ_Generation_2025-06-28_18_53_06Z-836062246.2A24_L001-ds.d8abe0c9b6764755a8b18cdb7c2b8a19.skinmicrobiome2_S2_L001 | MN | MN 24h | Arm | P2 | 2A24 | 2A24.skinmicrobiome2 |
| FASTQ_Generation_2025-06-28_18_53_06Z-836062246.2Aswab_L001-ds.778e158273d74ff99606dbebabf0c2b8.skinmicrobiome3_S3_L001 | swab | Swab | Arm | P2 | 2Aswab | 2Aswab.skinmicrobiome3 |
| FASTQ_Generation_2025-06-28_18_53_06Z-836062246.2B20_L001-ds.dd06a933297d44ecae51687c5793b50a.skinmicrobiome4_S4_L001 | MN | MN 20 min | Back | P2 | 2B20 | 2B20.skinmicrobiome4 |
| FASTQ_Generation_2025-06-28_18_53_06Z-836062246.2B24_L001-ds.ab228dda0767494ab832871519416537.skinmicrobiome5_S5_L001 | MN | MN 24h | Back | P2 | 2B24 | 2B24.skinmicrobiome5 |
| FASTQ_Generation_2025-06-28_18_53_06Z-836062246.2Bswab_L001-ds.aa622fe75a9045cbb42d9915e6e9308f.skinmicrobiome6_S6_L001 | swab | Swab | Back | P2 | 2Bswab | 2Bswab.skinmicrobiome6 |
| FASTQ_Generation_2025-06-28_18_53_06Z-836062246.2airswab_L001-ds.b3c24d4a57e243b293bfd7b77ccb2ef3.skinmicrobiome7_S7_L001 | air | Air | Air | P2 | 2airswab | 2airswab.skinmicrobiome7 |
| FASTQ_Generation_2025-06-28_18_53_06Z-836062246.4A20_L001-ds.f071e6d5d7904818978c43ca0fffc986.skinmicrobiome8_S8_L001 | MN | MN 20 min | Arm | P4 | 4A20 | 4A20.skinmicrobiome8 |
| FASTQ_Generation_2025-06-28_18_53_06Z-836062246.4A24_L001-ds.09300f983fde492bbbaf6ddb7c8b4a30.skinmicrobiome9_S9_L001 | MN | MN 24h | Arm | P4 | 4A24 | 4A24.skinmicrobiome9 |
| FASTQ_Generation_2025-06-28_18_53_06Z-836062246.4Aswab_L001-ds.78edc06f81ae498abcdd041ecc995d4d.skinmicrobiome10_S10_L001 | swab | Swab | Arm | P4 | 4Aswab | 4Aswab.skinmicrobiome10 |
| FASTQ_Generation_2025-06-28_18_53_06Z-836062246.4B20_L001-ds.373b1408ce614599bec43074764e0554.skinmicrobiome11_S11_L001 | MN | MN 20 min | Back | P4 | 4B20 | 4B20.skinmicrobiome11 |
| FASTQ_Generation_2025-06-28_18_53_06Z-836062246.4B24_L001-ds.33ee25e949314207ab9620a1e2364cc3.skinmicrobiome12_S12_L001 | MN | MN 24h | Back | P4 | 4B24 | 4B24.skinmicrobiome12 |
| FASTQ_Generation_2025-06-28_18_53_06Z-836062246.4Bswab_L001-ds.84b3e11b08ae4be6ba7d29352882dff3.skinmicrobiome13_S13_L001 | swab | Swab | Back | P4 | 4Bswab | 4Bswab.skinmicrobiome13 |
| FASTQ_Generation_2025-06-28_18_53_06Z-836062246.4airswab_L001-ds.2d2a9116d8a542029df2c50a238dd465.skinmicrobiome14_S14_L001 | air | Air | Air | P4 | 4airswab | 4airswab.skinmicrobiome14 |
| FASTQ_Generation_2025-06-28_18_53_06Z-836062246.6A20_L001-ds.d70d577d27524fd8922636d56d0af1ce.skinmicrobiome15_S15_L001 | MN | MN 20 min | Arm | P6 | 6A20 | 6A20.skinmicrobiome15 |
| FASTQ_Generation_2025-06-28_18_53_06Z-836062246.6A24_L001-ds.4da0206162cb4901851e8d20f75b7b31.skinmicrobiome16_S16_L001 | MN | MN 24h | Arm | P6 | 6A24 | 6A24.skinmicrobiome16 |
| FASTQ_Generation_2025-06-28_18_53_06Z-836062246.6Aswab_L001-ds.2c2be108ef0542669e1352e6c15d034a.skinmicrobiome17_S17_L001 | swab | Swab | Arm | P6 | 6Aswab | 6Aswab.skinmicrobiome17 |

| | | | | | | |
|---|---|---|---|---|---|---|
| FASTQ_Generation_2025-06-28_18_53_06Z-836062246.6B20_L001-ds.8f3c08124d4242e7ba21fbdda7597d3d.skinmicrobiome18_S18_L001 | MN | MN 20 min | Back | P6 | 6B20 | 6B20.skinmicrobiome18 |
| FASTQ_Generation_2025-06-28_18_53_06Z-836062246.6B24_L001-ds.0c2c3347d6d94b5293865357be768fd6.skinmicrobiome19_S19_L001 | MN | MN 24h | Back | P6 | 6B24 | 6B24.skinmicrobiome19 |
| FASTQ_Generation_2025-06-28_18_53_06Z-836062246.6Bswab_L001-ds.0e1041c88b8149749623d21090f260e8.skinmicrobiome20_S20_L001 | swab | Swab | Back | P6 | 6Bswab | 6Bswab.skinmicrobiome20 |
| FASTQ_Generation_2025-06-28_18_53_06Z-836062246.6airswab_L001-ds.d52142893d6743c6be7209b43ca23f4f.skinmicrobiome21_S21_L001 | air | Air | Air | P6 | 6airswab | 6airswab.skinmicrobiome21 |
| FASTQ_Generation_2025-06-28_18_53_06Z-836062246.7A20_L001-ds.fd5835c6e80b47a5a3a1928b00908877.skinmicrobiome22_S22_L001 | MN | MN 20 min | Arm | P7 | 7A20 | 7A20.skinmicrobiome22 |
| FASTQ_Generation_2025-06-28_18_53_06Z-836062246.7A24_L001-ds.928401711ab14508a4f4fda24cb15ee4.skinmicrobiome23_S23_L001 | MN | MN 24h | Arm | P7 | 7A24 | 7A24.skinmicrobiome23 |
| FASTQ_Generation_2025-06-28_18_53_06Z-836062246.7Aswab_L001-ds.4db6d7564706423eba002ba03a2f54fb.skinmicrobiome24_S24_L001 | swab | Swab | Arm | P7 | 7Aswab | 7Aswab.skinmicrobiome24 |
| FASTQ_Generation_2025-06-28_18_53_06Z-836062246.7B20_L001-ds.68f74a9ea563411ca5d00d2a9d01fcaa.skinmicrobiome25_S25_L001 | MN | MN 20 min | Back | P7 | 7B20 | 7B20.skinmicrobiome25 |
| FASTQ_Generation_2025-06-28_18_53_06Z-836062246.7B24_L001-ds.e6453947646f405280b7ccb462e05db6.skinmicrobiome26_S26_L001 | MN | MN 24h | Back | P7 | 7B24 | 7B24.skinmicrobiome26 |
| FASTQ_Generation_2025-06-28_18_53_06Z-836062246.7airswab_L001-ds.6d6bb513f5db4d4f8da69c8d8e689322.skinmicrobiome28_S28_L001 | air | Air | Air | P7 | 7airswab | 7airswab.skinmicrobiome28 |
| FASTQ_Generation_2025-06-28_18_53_06Z-836062246.8A20_L001-ds.4dd954c6006b4f0686e2cd2e656613c5.skinmicrobiome29_S29_L001 | MN | MN 20 min | Arm | P8 | 8A20 | 8A20.skinmicrobiome29 |
| FASTQ_Generation_2025-06-28_18_53_06Z-836062246.8A24_L001-ds.5c312b9ceafa47e89710f363df4f868a.skinmicrobiome30_S30_L001 | MN | MN 24h | Arm | P8 | 8A24 | 8A24.skinmicrobiome30 |
| FASTQ_Generation_2025-06-28_18_53_06Z-836062246.8Aswab_L001-ds.d82fa4befb554abda2338820ea16b629.skinmicrobiome31_S31_L001 | swab | Swab | Arm | P8 | 8Aswab | 8Aswab.skinmicrobiome31 |
| FASTQ_Generation_2025-06-28_18_53_06Z-836062246.8B20_L001-ds.b9cebabcc6224a088e322c57cbc8213c.skinmicrobiome32_S32_L001 | MN | MN 20 min | Back | P8 | 8B20 | 8B20.skinmicrobiome32 |
| FASTQ_Generation_2025-06-28_18_53_06Z-836062246.8B24_L001-ds.4302d1f7062f41b6b813edecf7eb9fef.skinmicrobiome33_S33_L001 | MN | MN 24h | Back | P8 | 8B24 | 8B24.skinmicrobiome33 |
| FASTQ_Generation_2025-06-28_18_53_06Z-836062246.8Bswab_L001-ds.3bf52a3f0ea247c5b378ea8419b44327.skinmicrobiome34_S34_L001 | swab | Swab | Back | P8 | 8Bswab | 8Bswab.skinmicrobiome34 |
| FASTQ_Generation_2025-06-28_18_53_06Z-836062246.8airswab_L001-ds.123815a5d8264a83b3923c87b60459cf.skinmicrobiome35_S35_L001 | air | Air | Air | P8 | 8airswab | 8airswab.skinmicrobiome35 |
| N/A | Alginate | Control MN | CTRL | Control MN | Control MN | Alginate.HUskinmicrobiome1 |
| N/A | Blank | CTRL | CTRL | CTRL | RID001578Blank | RID001578Blank.RID001578Blank |

| | | | | | | |
|---|---|---|---|---|---|---|
| N/A | Extraction.control | Extraction.control | Air | CTRL | airnew | airnew.HUskinmicrobiome3 |
| N/A | Extraction.control | Extraction.control | Air | CTRL | Extraction neg. control | airold.HUskinmicrobiome4 |
| N/A | Non.Alginate | Alg.CTRL | CTRL | CTRL | nonalginate | nonalginate.HUskinmicrobiome2 |

### Supplementary Figures

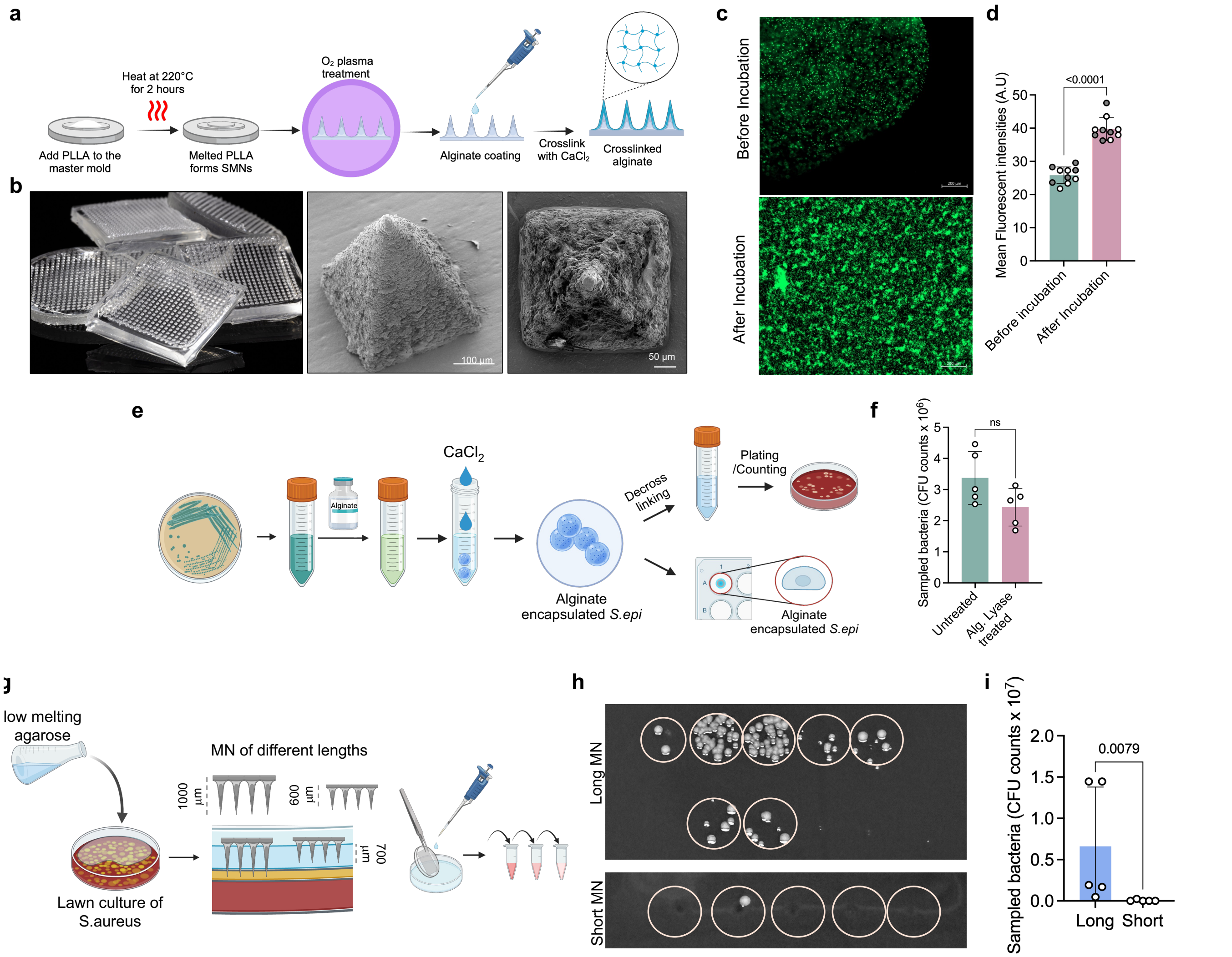


**Figure S1. Fabrication, structural characterization, and microbial viability validation of hydrogel-coated microneedle patches. a**, Schematic representation of the MN patch fabrication process from melt molding of PLLA to the hydrogel coating and crosslinking. **b**, Photograph showing the hydrogel coated MN patch and scanning electron micrographs showing the uncoated (left) and hydrogel coated (right) microneedle. **c**, Representative fluorescence micrographs showing GFP containing *S. epidermidis* encapsulated SLG20 hydrogel before and after overnight incubation to assess bacterial viability in alginate. **d**, Comparison of mean fluorescence intensities of the micrographs shown in c (n=10 data points per group). Grey and white dots represent the data from 2 iterations of experiments. **e**, Schematic view of assessment of bacterial viability after alginate decrosslinking using alginate lyase enzyme. **f**, comparison of CFU counts of bacteria before and after alginate lyase treatment (n=5 replicates per group). **g**, schematic representation of the bacterial sampling from different depths using the MN patches. **h**, Colony counts of the serial dilution experiments to quantify the sampled bacteria using short and long MN patches. **i**, CFU counts of bacteria sampled by the long and short MN patches (n=5 replicates per group). Data shown are mean±SEM. P values were determined using one-way ANOVA followed by Tukey's multiple comparisons test. Panel a, e, and g elements were created using BioRender.

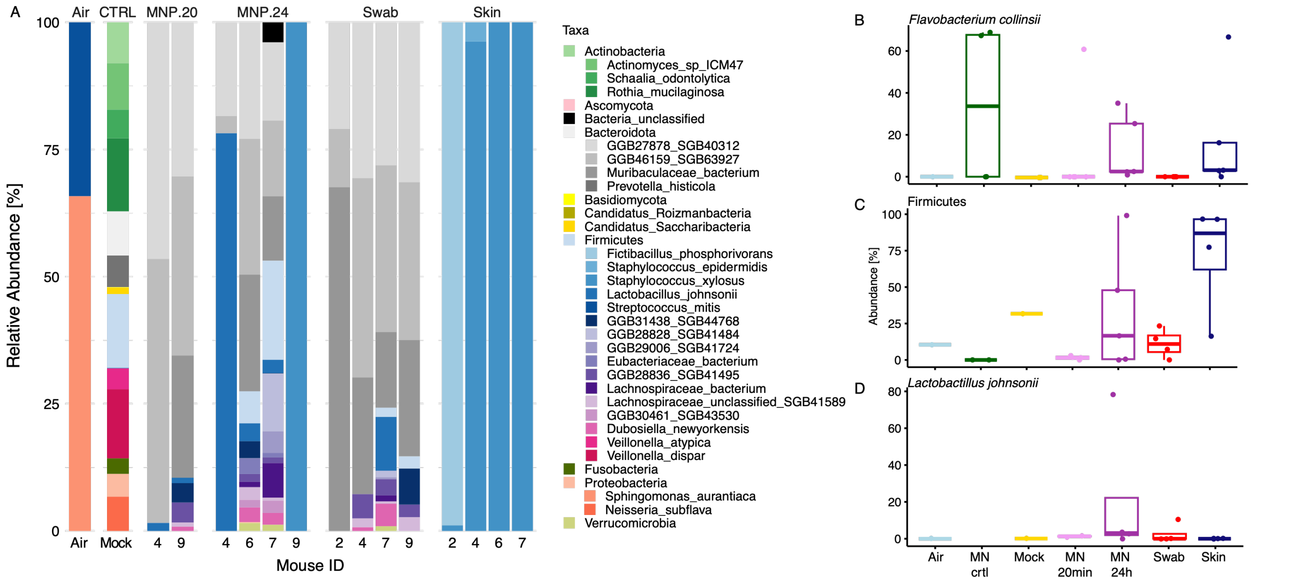


**Figure S2. Murine microbiome profiling and taxonomic composition across sampling modalities.** **a**, Relative abundance plots of the 25 abundant taxa are shown for all controls, 20 minute and 24 hour MN patches as well as skin swabs and skin tissues. Remaining species were grouped by their respective phylum. Taxa of interest such as the (**b**) *Flavobacterium* contaminant and (**c**) the Firmicutes phylum and (**d**) *Lactobacillus johnsonii* are shown separately.

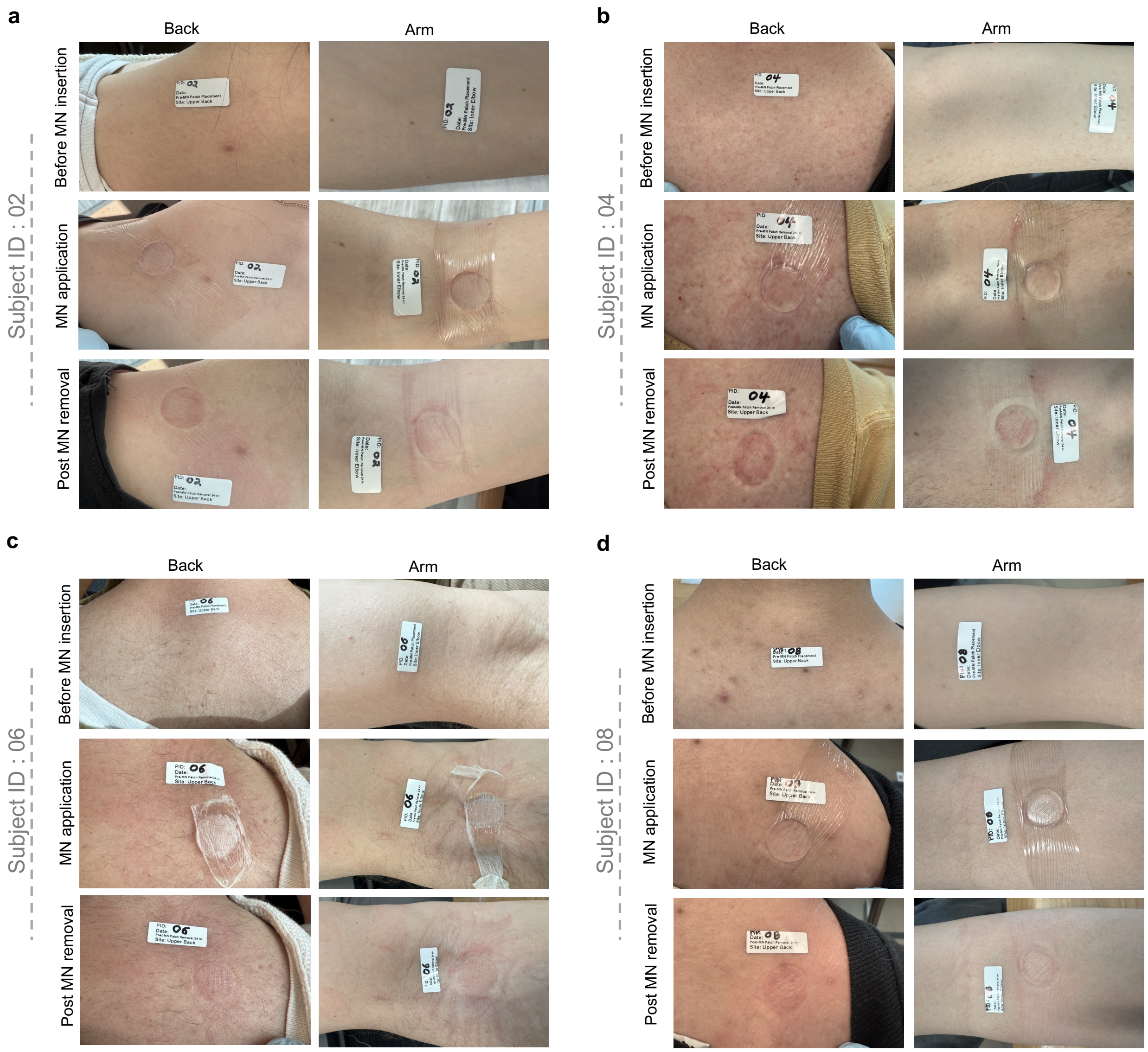


**Figure S3. Representative images of microneedle patch application in human subjects.** Photographs of pre- and post-MN patch application on the arm and back of human volunteers with subject IDs 02 (**a**), 04 (**b**), 06 (**c**), and 08 (**d**).

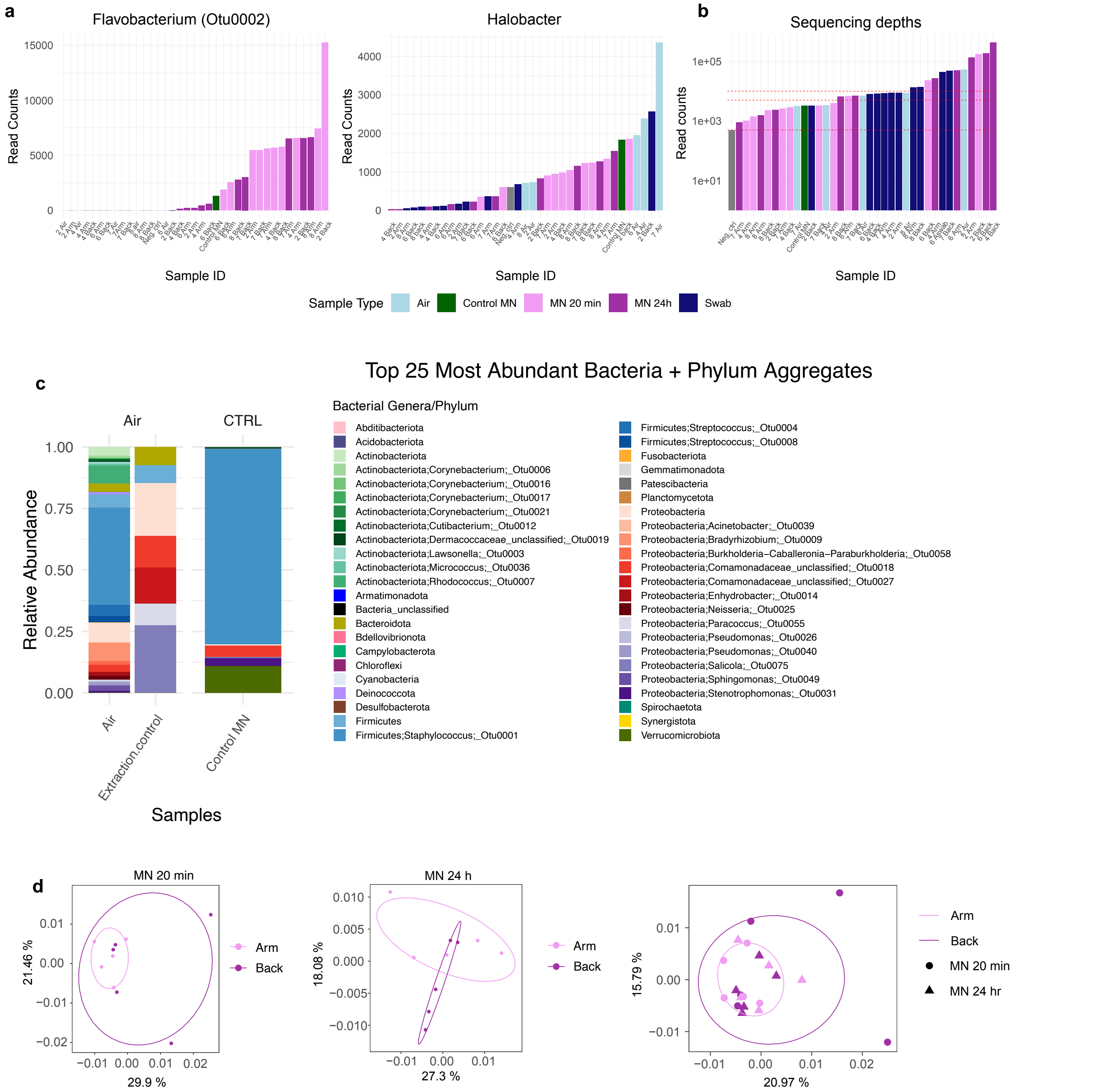


**Figure S4. Quality control and contaminant filtering of 16S sequencing datasets.** Quality control of 16S RNA gene sequencing data. **a**, read counts per sample of Flavobacterium (OTU2) per sample, a contaminant from the alginate lyase used during MN patch preparation and Halobacterium, an archaea over amplified during PCR. **b**, Read counts per sample after removal of contaminants (OTU2) and Halobacter with red dashed lines indicating 500, 5000 and 10000 reads. **c**, Abundance plot of top 25 genera and all phyla (after removal of OTU2, and Halobacter OTUs) obtained from all 16S sequenced samples including technical controls from MN patch preparation (Alginate = unused alginate-treated; nonalginate = not-alginate-treated MNs) and skin sampling controls (air swabs collected during patient sampling ("air") and air swabs (extraction control) collected during DNA extraction (airold). **d**, PCOA (based on Bray-Curtis dissimilarity index) showing the MN samples collected from arms and the backs of individuals.

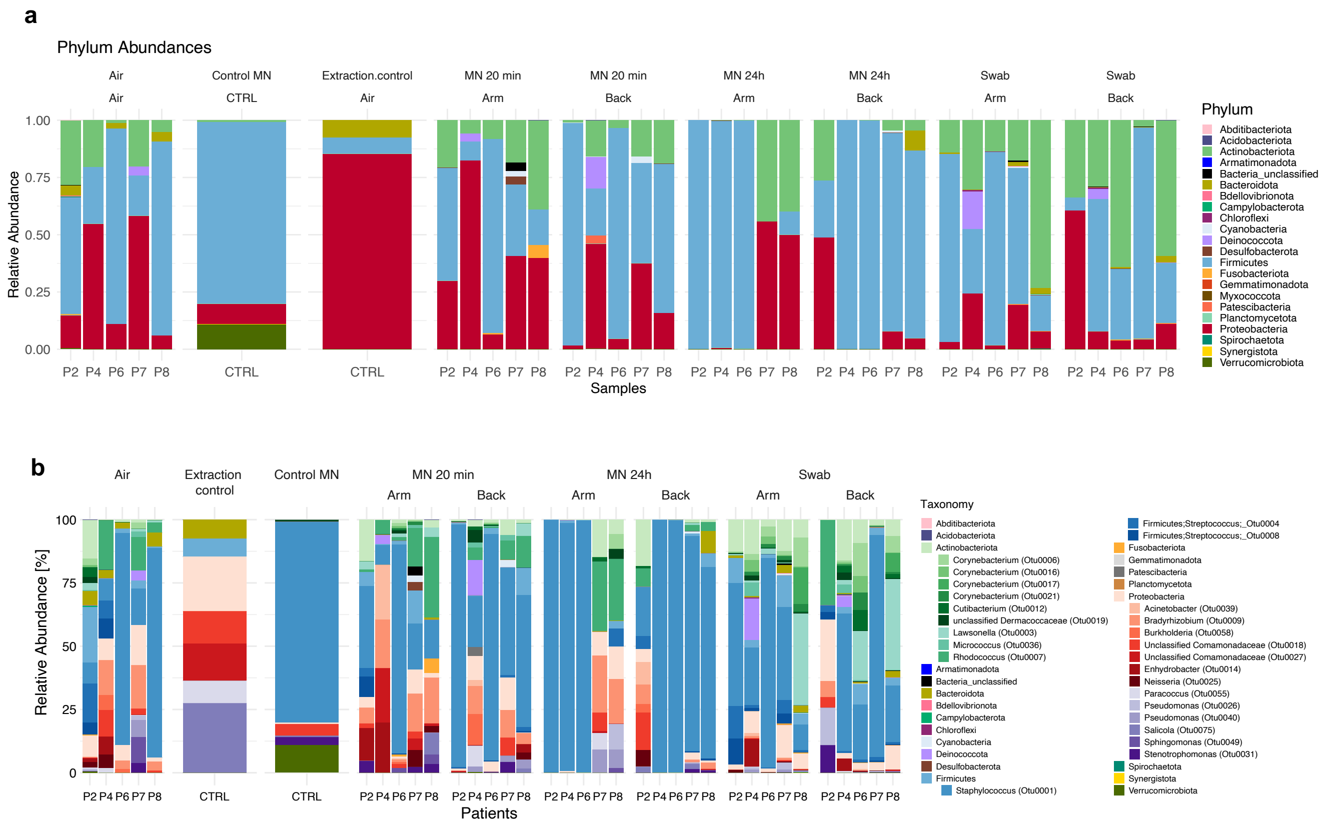


**Figure S5. Microbial composition of human skin samples and technical controls.** **a**, Relative abundance plot of all phyla (after removal of contaminants) obtained from all 16S sequenced samples including technical controls from MN patch fabrication. **b**, Relative abundance plots of the 25 abundant taxa are shown for all controls, 20 min and 24 hour MN patches as well as skin swabs for both arm and back of all human subjects. Remaining species were grouped by their respective phylum.

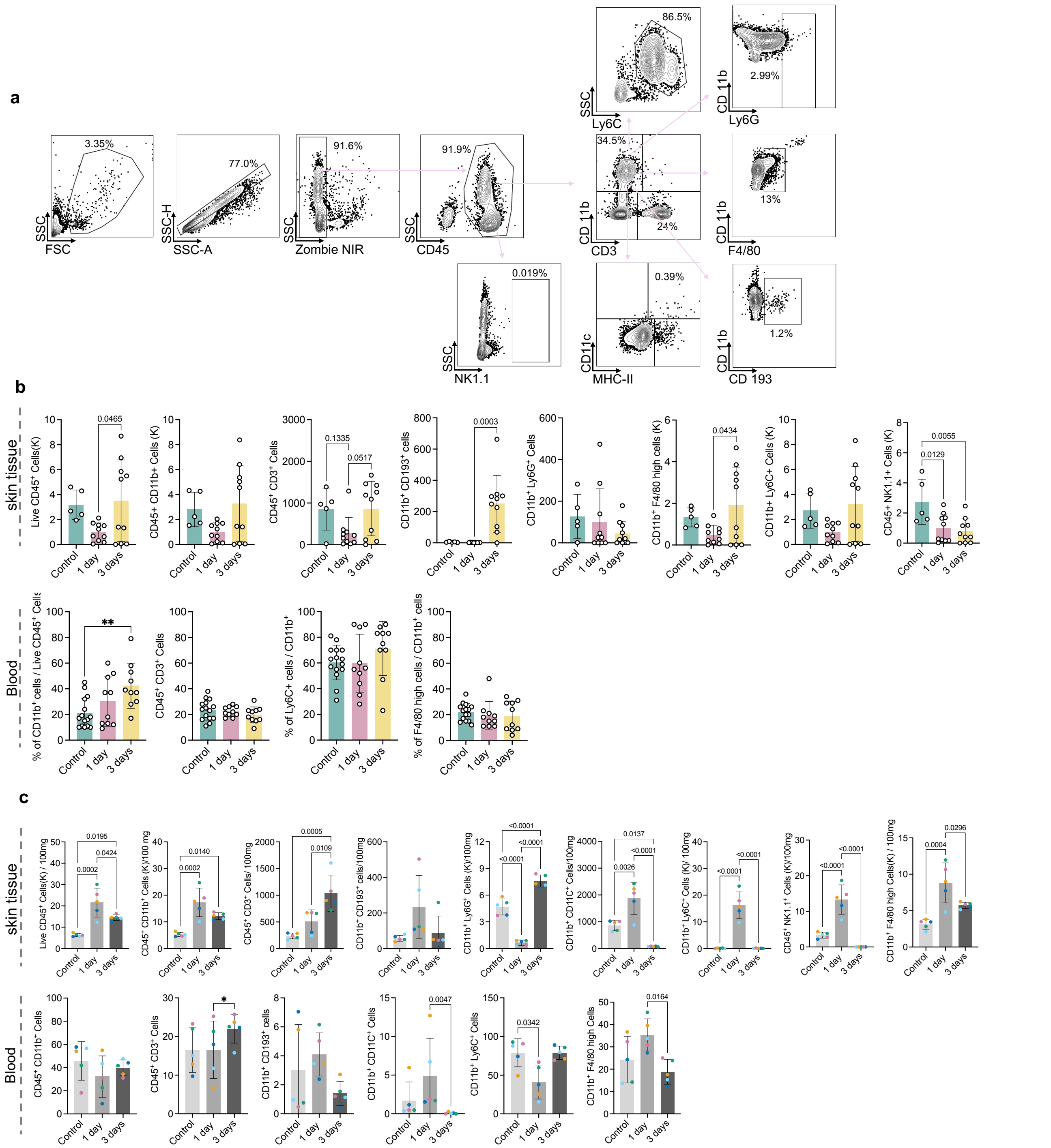


**Figure S6. Flow cytometry gating strategy and validation of immune cell dynamics in colonization models. a**, Representative flow cytometry plots demonstrate gating strategies for identifying single cell population of live leukocytes, T cells, myeloid cells, NK cells, dendritic cells, eosinophils, macrophages, neutrophils and monocyte subsets. **b**, Enumeration of live immune cells from mice skin tissue and blood after *S. aureus* colonization. n=10 mice in all groups. **c**, Immune cell counts from skin tissue and blood of mice after co-colonization with *S. epidermidis* and *S. aureus*. (panel c: each colored dot represents an individual mouse, with corresponding MN bacterial burden and immune cell measurements in Fig. 4f,g). n=5 mice in all groups. Data shown are mean±SEM. P values were determined using one-way ANOVA followed by Tukey's multiple comparisons test.
